# Cortical network mechanisms in subcallosal cingulate deep brain stimulation for depression

**DOI:** 10.1101/2023.10.31.23297406

**Authors:** M Scherer, IE Harmsen, N Samuel, GJB Elias, J Germann, A Boutet, CE MacLeod, P Giacobbe, NC Rowland, AM Lozano, L Milosevic

## Abstract

Identifying functional biomarkers of clinical success can contribute to therapy optimization, and provide insights into the pathophysiology of treatment-resistant depression and mechanisms underlying the potential restorative effects of subcallosal cingulate deep brain stimulation.

Magnetoencephalography data were obtained from 15 individuals who underwent subcallosal cingulate deep brain stimulation for treatment-resistant depression and 25 healthy subjects. The first objective herein was to identify region-specific oscillatory modulations for the identification of discriminative network nodes expressing (i) pathological differences in TRD (responders and non-responders, stimulation-OFF) compared to healthy subjects, which (ii) were counteracted by stimulation in a responder-specific manner. The second objective of this work was to further explore the mechanistic effects of stimulation intensity and frequency.

Oscillatory power analyses led to the identification of discriminative regions that differentiated responders from non-responders based on modulations of increased alpha (8-12 Hz) and decreased gamma (32-116 Hz) power within nodes of the default mode, central executive, and somatomotor networks, Broca’s area, and lingual gyrus. Within these nodes, it was also found that low stimulation frequency had stronger effects on oscillatory modulation than increased stimulation intensity.

The identified discriminative network profile implies modulation of pathological activities in brain regions involved in emotional control/processing, motor control, and the interaction between speech, vision, and memory, which have all been implicated in depression. This modulated network profile may represent a functional substrate for therapy optimization. Stimulation parameter analyses revealed that oscillatory modulations can be strengthened by increasing stimulation intensity or, to an even greater extent, by reducing frequency.

## Introduction

Clinical depression (major depressive disorder) affects 6 % of the adult population annually (1), and up to 30 % of affected individuals are treatment-resistant (2). Treatment-resistant depression (TRD) can be characterized by the insufficient response to conventional treatments, which typically entails the combination of antidepressant medication, psychotherapy, and electroconvulsive therapy (3). As such, TRD represents a major social and economic burden worldwide (with affected individuals experiencing social and occupational dysfunction, and poor physical health, causing increased healthcare utilization), prompting research in the development of novel therapies for TRD.

Neuromodulation modalities, such as repetitive transcranial magnetic stimulation (rTMS), transcranial direct current stimulation (tDCS), and deep brain stimulation (DBS), are being researched as potential treatments for TRD. Of these interventions, DBS for TRD enables the direct stimulation of subcortical regions that cannot be targeted by non-invasive methods and has the additional advantage that it may provide sustained therapeutic effects without the requirement of frequent clinical visits for therapy administration. DBS appears safe for TRD when targeting the subcallosal cingulate (SCC), medial forebrain bundle, or ventral capsule/striatum areas. However, data on the long-term efficacy of the treatment are highly variable. After two or more years of stimulation, the treatment response varied between 23.3-71.4 % with striatal/capsular area DBS (n = 48) and 25.0-64.7 % with SCC-DBS (n = 105) (4–7).

While DBS efficacy can be quickly evaluated in people with Parkinson’s disease or essential tremor due to the near instantaneous relief of motor symptoms (8), this does not apply to psychological disorders such as TRD. In this population, clinical evaluations are typically performed retrospectively using clinical assessment batteries which average mood over the course of weeks or months; hence the evaluation of multiple DBS configurations, the stimulation titration process, extends over months or years (9). In addition to the immense time requirement, this process is also be subject to a high degree of external confounding influences (i.e., job loss/stress). Two contributors critically affecting outcome following the surgical implantation of the DBS leads are contact selection and stimulation programming. Given the high cost (in both patient well-being and healthcare resources) of qualitatively assessing the efficacy of a single stimulation configuration (contact selection and stimulation programming), appropriate early indicators of treatment success are strongly warranted by patients and healthcare professionals.

The first objective of this work was to use magnetoencephalography (MEG) to identify electrophysiological characteristics supportive in DBS lead placement confirmation and DBS contact selection. Therefore, we sought to identify a “minimal discriminative functional network response profile” that could differentiate treatment responders from non-responders when stimulated at a given DBS contact. As individually optimized settings are not yet established in new patients, the discriminative response profile would have to be inducible through generic, uniform stimulation parameters.

The second objective of this work was to gain a better mechanistic understanding of the impacts of stimulation intensity and frequency on the modulation of cortical network activity. Hence, we investigated the contrast of high versus low stimulation intensity (constraining frequency) and high versus low frequency (constraining intensity). In clinical settings, stimulation intensity is often titrated in accordance to side effect profiles (10); however, investigations of frequency-dependence are of particular interest as many new experimental DBS indications default to the use of 130 Hz stimulation due to its success in movement disorders, rather than a comprehensive understanding of the functional implications.

## Material and methods

### Participants

People with TRD who had previously been implanted with bilateral SCC-DBS electrodes (n = 15; Figure 1 A) were included in this study (details about surgical procedures are provided within the Supplementary Materials). Age- and sex-matched healthy controls (HC; n = 25) without major neurological or psychiatric diseases were also recruited. All subjects provided written informed consent. The study was approved by the University Health Network Research Ethics Board (Toronto, Canada) and was conducted in accordance with the Declaration of Helsinki.

**Figure 1.**
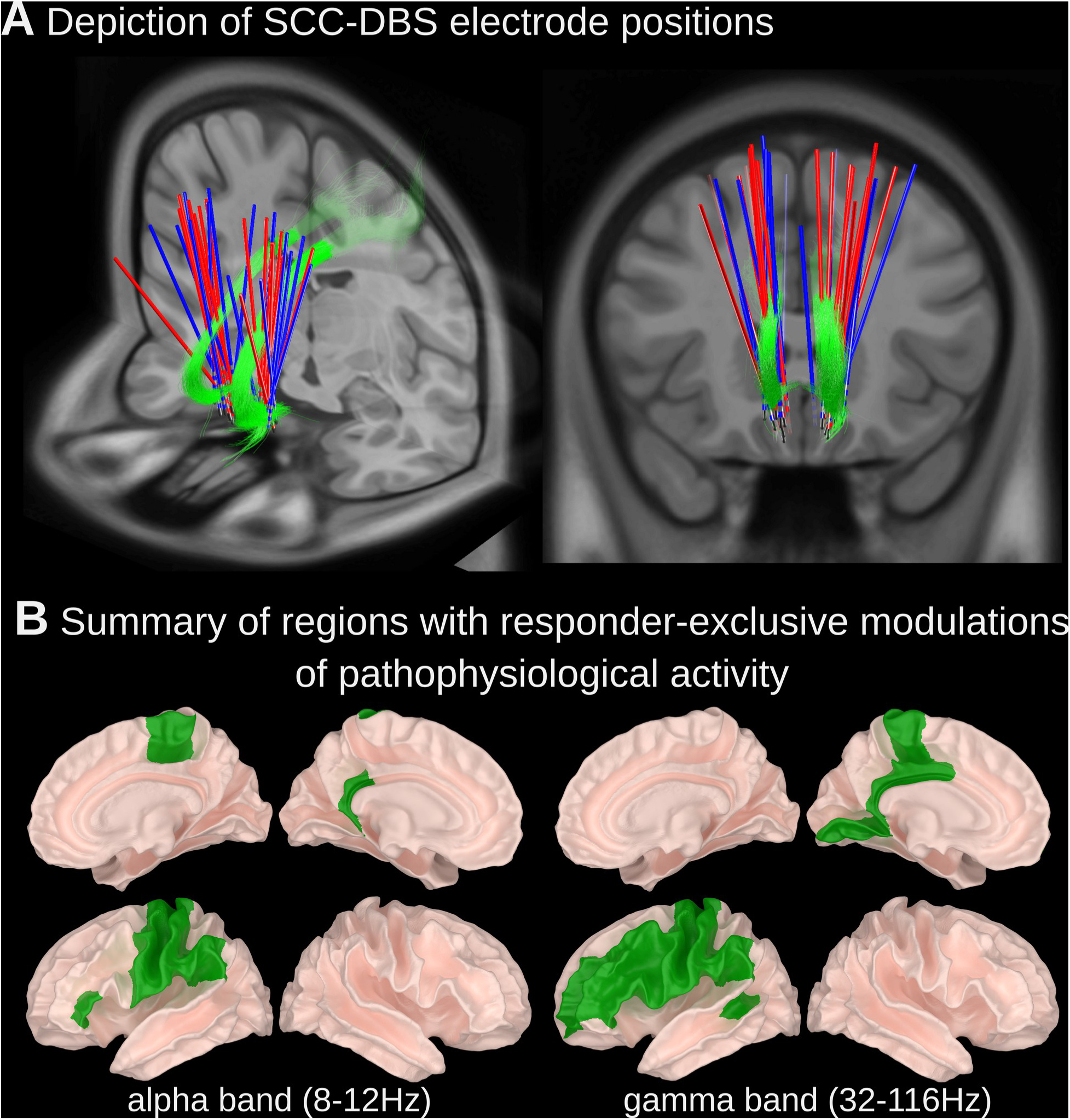
(A) Depiction of SCC-DBS electrode positions and (B) summary of regions with responder-specific modulations of pathophysiological activity. (A) DBS lead positions are shown for responder (blue) and non-responder (red) subgroups relative to the cingulum bundle (green). Lead localization was done using LeadDBS (detailed methods available within Supplementary Material). (B) Brain regions that (1) expressed pathological changes in oscillatory power in people with depression (SCC-DBS OFF) compared to HCs, and (2) SCC-DBS ON counteracted these pathological changes.

For people with TRD, disease severity was measured preoperatively and 12 months postoperatively by a psychiatrist (P.G.) using the 17-item Hamilton Depression Rating Scale (HAM-D17; Table 1). Seven patients were considered treatment responders (HAM-D17 change ≥ 50%), and eight were considered non-responders. Clinical outcomes of patients 3, 6, 9, and 14 were reported in previous studies (11, 12). The patient’s clinical details and therapeutic DBS settings are listed in Table 1. Additional clinical history data is available in Supplementary Table 1.

**Table 1.** Clinical characteristics and therapeutic DBS settings of TRD participants.

| Subject | Age of MDD onset | MDD duration (y) | MDD features |  | DBS duration (y) | HAMD-17 score |  | % Improvement with SCC-DBS (%)<sup>a</sup> | Responder status | Therapeutic DBS settings |  |  |  |
| --- | --- | --- | --- | --- | --- | --- | --- | --- | --- | --- | --- | --- | --- |
|  |  |  | Melancholic | Anxious |  | SCC-DBS OFF | SCC-DBS ON |  |  | Bilateral contact configuration | Amplitude (mA) | Pulse width (µs) | Frequency (Hz) |
|  |  |  |  |  |  | *Pre-operative score* | *12-month post-operative score* |  |  |  |  |  |  |
| 1 | 20s | 0-5 | no | yes | <1 | 20-30 | 0-10 | 80-90 | Responder | C+1- | 3.5 | 91 | 130 |
| 2 | 30s | 20-30 | yes | yes | <1 | 30-40 | 10-20 | 50-60 | Responder | C+1- | 4.5 | 91 | 130 |
| 3 | 20s | 20-30 | no | yes | 5-10 | 30-40 | 10-20 | 50-60 | Responder | * | * | * | * |
| 4 | 30s | 10-20 | yes | yes | 1-5 | 30-40 | 10-20 | 50-60 | Responder | C+1-2- | 6.5 | 91 | 130 |
| 5 | 10s | 30-40 | yes | yes | <1 | 20-30 | 10-20 | 30-40 | Non-responder | C+1- | 5.5 | 91 | 130 |
| 6 | 10s | 30-40 | yes | no | 10-15 | 20-30 | 10-20 | 50-60 | Responder | C+1- | 5.5 | 60 | 130 |
| 7 | 10s | 20-30 | yes | no | <1 | 20-30 | 20-30 | -10-0 | Non-responder | C+1-2- | 4.5 | 91 | 130 |
| 8 | 10s | 30-40 | no | yes | 5-10 | 20-30 | 10-20 | 40-50 | Non-responder | C+2- | 9 | 87 | 130 |
| 9 | 10s | 30-40 | yes | yes | 5-10 | 20-30 | 10-20 | 10-20 | Non-responder | C+1- | 6 | 90 | 130 |
| 10 | 20s | 20-30 | no | yes | 1-5 | 20-30 | 20-30 | -10-0 | Non-responder | 1+/2- | 7.5 | 91 | 130 |
| 11 | 10s | 30-40 | yes | no | 1-5 | 20-30 | 20-30 | -10-0 | Non-responder | C+3- | 7.5 | 91 | 130 |
| 12 | 10s | 10-20 | no | yes | 1-5 | 20-30 | 0-10 | 70-80 | Responder | C+2- | 11 | 91 | 130 |
| 13 | 20s | 10-20 | no | yes | 1-5 | 20-30 | 0-10 | 70-80 | Responder | C+1- | 7 | 91 | 130 |
| 14 | 30s | 20-30 | * | * | 5-10 | 20-30 | 10-20 | 40-50 | Non-responder | C+1- | 7 | 87 | 130 |
| 15 | 10s | 20-30 | yes | yes | 1-5 | 20-30 | 20-30 | 10-20 | Non-responder | C+1-2- | 7 | 91 | 130 |

\*Depressive symptom improvement represents the percent change in HAMD-17 score between SCC-DBS OFF (pre-operative score) and ON (12-month post-operative score). Percent improvement is calculated as [(DBS ON - DBS OFF)/DBS OFF] x (-100). Positive values represent an improvement in symptoms (lower HAMD-17 score) with SCC-DBS; negative values represent a worsening in symptoms (higher HAMD-17 score) with SCC-DBS. \*\*60 µs pulse width used for patient 6. DBS, deep brain stimulation; HAMD-17, 17-item Hamilton Depression Rating Scale; SCC, subcallosal cingulate

### MEG recordings

The first objective of this study was to identify a minimal discriminative functional network profile that could differentiate responders from non-responders. MEG data (306 channels each sampled at 1,000 Hz) were acquired (resting-state, supine-positioned, eyes-closed) from HCs and people with TRD during DBS OFF and DBS ON (three minutes per condition). In trials with DBS, stimulation was applied bilaterally at 130 Hz, 1.5 mA, 90 µs pulse width, bipolar 1-2+. Instead of using variable/patient-specific DBS parameters (see Table 1 for clinical DBS parameters), we used a unified DBS parameter set across all patients to establish a standardized approach for deriving a “minimal” discriminative functional network response profile that may be predictive of therapeutic success in prospective contexts (i.e., when optimized clinical parameters are otherwise unknown in the early stages of therapy initiation).

The second objective of this study was to investigate how changes to stimulation settings may impact cortical network activity. To do this, three intensity/frequency combinations of stimulation were applied unilaterally in the left hemisphere: 1.5 mA & 130 Hz (baseline condition); 3.0 mA & 130 Hz (to study the effect of stimulation intensity); and 1.5 mA & 20 Hz (to study the effect of stimulation frequency).

For analyses, after digitally removing ambient artifacts and high-frequency components (>240 Hz), data were downsampled to 120 Hz. MEG data were then transformed from sensor to source space and clustered within anatomical boundaries defined in the Desikan-Killiany atlas (Supplementary Figure 1) (13). Using representative sub-bands, cluster-wise normalized spectral power was calculated using Welch’s method for delta (2-4 Hz), theta (4-8 Hz), alpha (8-12 Hz), beta (12-32 Hz), and gamma (32-116 Hz) frequencies. Additional methodological details regarding source reconstruction and data processing are available in Supplementary Materials.

### Statistics

Effects were quantified using linear mixed models (Python-FiNNPy (14)/R-lme4 (15)). For the first objective of this work (derivation of a minimally discriminative functional network response profile) a cortical region was considered to be a discriminative node if found to: (1) express pathological activity in responders and non-responders with SCC-DBS OFF (compared to HCs) *and* (2) be modulated by SCC-DBS ON in a compensatory way in responders only. For condition 1, pathological activity was defined as a significant difference in spectral power between HC vs. TRD (responder and non-responder subgroups with SCC-DBS OFF). For condition 2, the significant difference had to be negated or reversed by SCC-DBS in responders while unchanged in non-responders. In other words, the pathological effect had to be modulated to a level statistically indistinguishable from HC (termed “normalized”) or to a level in the opposite direction to the pathological effect (termed “overcompensated”) in the responder subgroup, while being unmodulated in the non-responder subgroup.

For the second objective of this work (investigating the effects of changing stimulation settings), we applied the same statistical criteria at regions of interest identified as in the first part of the analysis. The validity and reliability of all observations were confirmed using both 15-second and 30-second windows for data epoching. Since the multi-layered statistical approach necessitates a feature to be significant across three different hypotheses times two epoch window sizes, a high threshold for statistical significance was enforced, and no further correction methods were employed. Statistical significance was set at p<0.05. Detailed statistical values are available in Supplementary Table 2.

### Data availability

The study data may be shared upon reasonable request to the corresponding author.

## Results

Resting-state magnetoencephalography (MEG) recordings were acquired from people with TRD (n = 15) during SCC-DBS OFF and ON and from HCs (n = 25). The characteristics of the TRD study participants are presented in Table 1. HCs comprised 12 males and 13 females, with an average age of 38.1 ± 3.9 years. There were no significant differences between HC and TRD group means for age (p = 0.179, t[38] = −1.37) or sex (p = 0.462, χ2[2,40] = 0.54).

Regions of interest were limited to those in which both of the following criteria were satisfied: (1) a statistical difference was identified in people with TRD (responders and non-responders with SCC-DBS OFF), which was (2) counteracted by SCC-DBS in responders only (“normalized” to a level statistically indistinguishable from HC or statistically “overcompensated” such that the effect was significant but in the opposite direction to the pathological effect). This data-driven approach led to identifying a discriminative network (summarized in Figure 1 B). Spatially, all identified effects were located within the left hemisphere, except for the right paracentral lobule. Spectrally, the identified effects were limited to the normalization of the increased alpha band (8-12 Hz) as well as the normalization and overcompensation of decreased gamma (32-116 Hz) power.

### Identification of a minimal discriminative functional network profile

#### Responder-specific normalization of increased alpha band power

In several cortical regions, we identified responder-specific normalizations of pathologically increased alpha band (8-12 Hz) power (Figure 2). Compared to HCs, TRD patients with SCC-DBS OFF had increased alpha activity, which was reduced to levels comparable to HCs in responders only when SCC-DBS was turned ON. This alpha normalization occurred in the left retrosplenial cortex, inferior frontal cortex pars triangularis, postcentral gyrus, precentral gyrus, supramarginal gyrus, and the right paracentral lobule.

**Figure 2.**
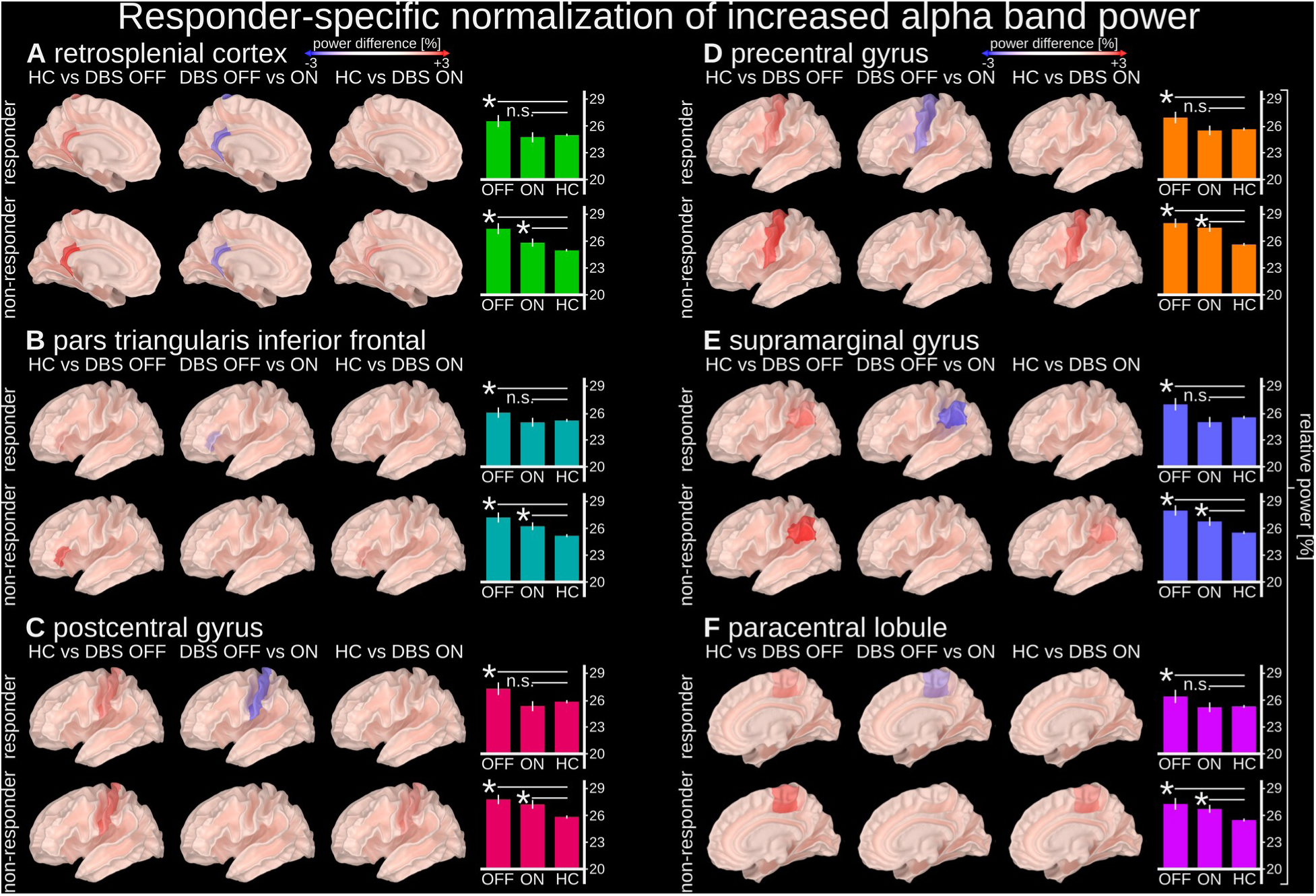
Responder-specific normalization of increased alpha power. Alpha power (8-12 Hz) in the highlighted regions was pathologically increased in people with depression (both responders and non-responders with SCC-DBS OFF) compared to HCs. The pathological increase in alpha power was normalized by SCC-DBS in responders specifically (i.e., statistically indistinguishable from HCs with SCC-DBS ON). Alpha power was measured as the ratio between the alpha (8-12 Hz) sub-band and broadband (2-116 Hz) power. Normalization of increased alpha power was observed in the left (A) retrosplenial cortex, (B) inferior frontal cortex, (C) postcentral gyrus, (D) precentral gyrus, (E) supramarginal gyrus, and (F) right paracentral lobule.

#### Responder-specific normalization of decreased gamma band power

We also identified responder-specific normalizations of pathologically decreased gamma band (32-116 Hz) power (Figure 3). Compared to HCs, TRD patients with SCC-DBS OFF had decreased gamma activity, which was increased to levels comparable to HCs in responders only when SCC-DBS was turned ON. This gamma normalization occurred in the left posterior cingulate, paracentral lobule, rostral middle frontal cortex (containing the dorsolateral prefrontal cortex; Figure 3 C), lingual gyrus, inferior frontal cortex pars opercularis, and the inferior frontal cortex pars triangularis.

**Figure 3.**
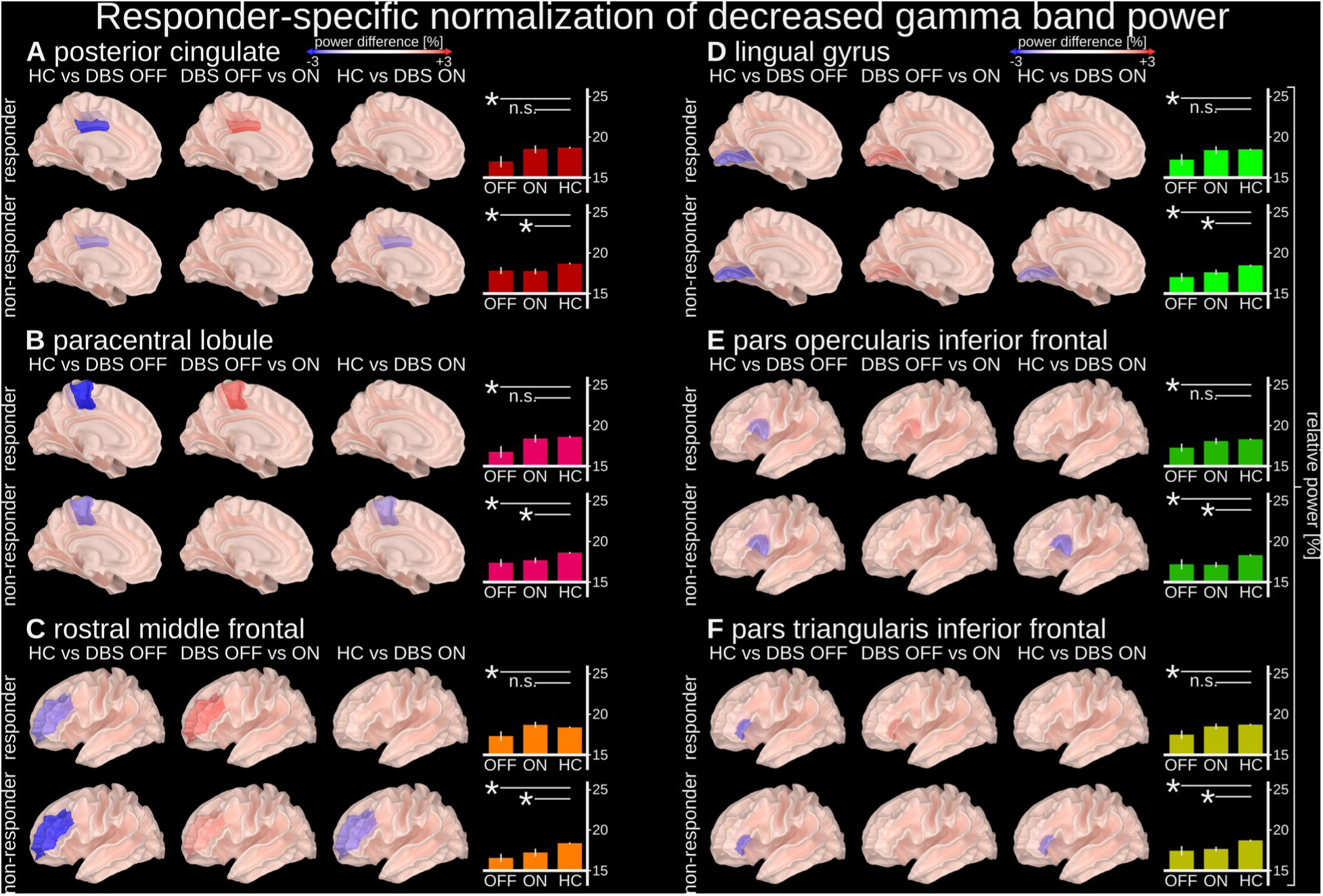
Responder-specific normalization of decreased gamma band power. Gamma power (32-116 Hz) in the highlighted regions was pathologically decreased in people with depression (both responders and non-responders with SCC-DBS OFF) compared to HCs. The pathological decrease in gamma power was normalized by SCC-DBS ON in responders specifically (i.e., statistically indistinguishable from HCs with SCC-DBS ON). Gamma power was measured as the ratio between the gamma (32-116 Hz) sub-band and broadband (2-116 Hz) power. Normalization of decreased gamma power was observed in the left (A) posterior cingulate, (B) paracentral lobule, (C) rostral middle frontal cortex, (D) lingual gyrus, (E) inferior frontal cortex pars opercularis, and (F) inferior frontal cortex pars triangularis.

#### Responder-specific overcompensation of decreased gamma band power

Lastly, we identified responder-specific overcompensation of pathologically decreased gamma band (32-116 Hz) power (Figure 4). Compared to HCs, TRD patients with SCC-DBS OFF had decreased gamma activity, which was increased beyond levels observed in HCs in responders only when SCC-DBS was turned ON. This gamma overcompensation occurred in the left retrosplenial cortex, precentral gyrus, supramarginal gyrus, postcentral gyrus, caudal middle frontal gyrus, and the banks of the superior temporal sulcus.

**Figure 4.**
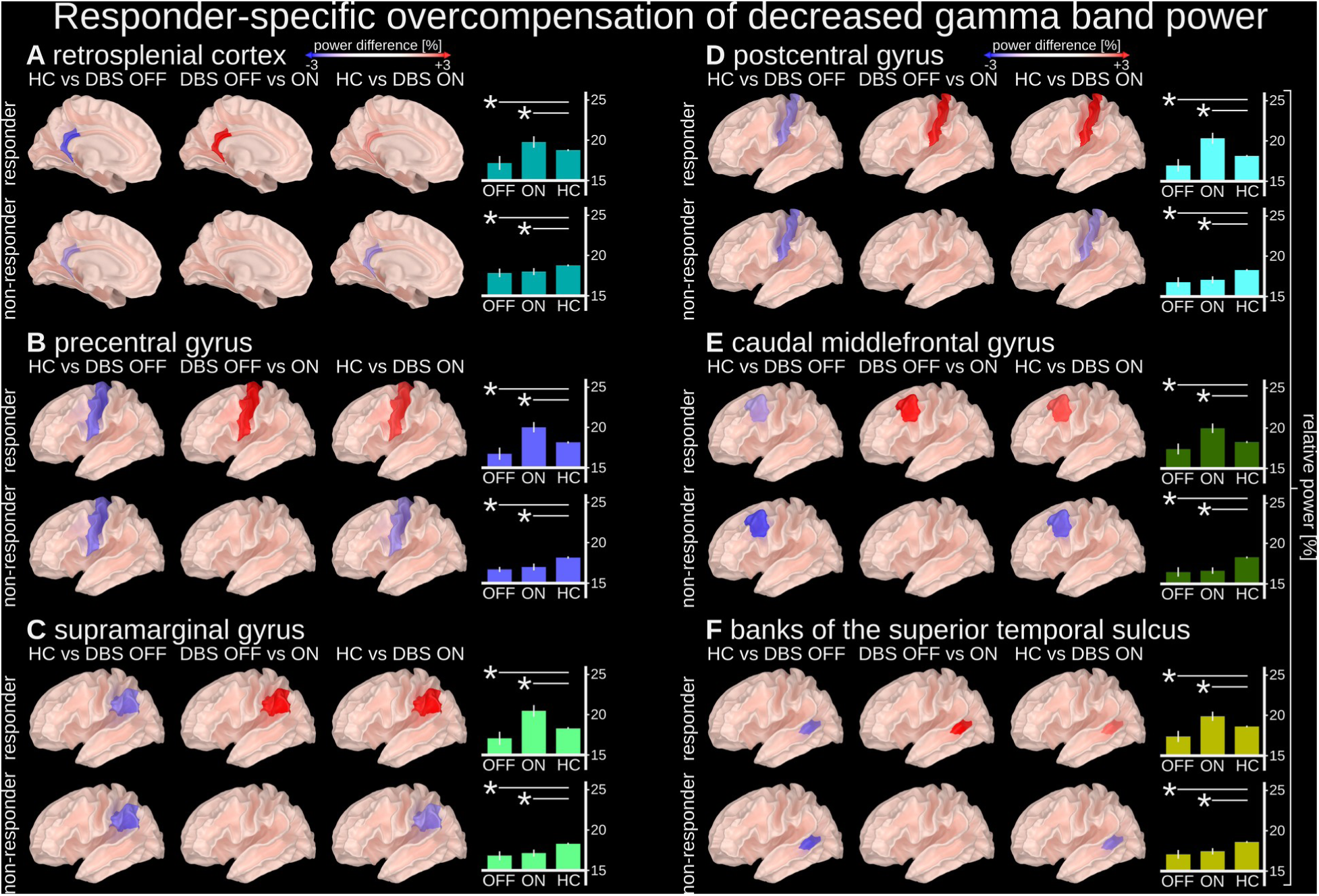
Responder-specific overcompensation of decreased gamma band power. Gamma activity (32-116 Hz) in the highlighted regions was found to be pathologically decreased in people with depression (both responders and non-responders with DBS OFF), but reversed by SCC-DBS ON in responders only (i.e., hypoactivity became hyperactivity). Gamma activity was measured as the ratio between a gamma (32-116 Hz) sub-band and broadband (2-116 Hz) activity. Overcompensation of gamma band hypoactivity was observed in the left (A) retrosplenial cortex, (B) precentral gyrus, (C) supramarginal gyrus, (D) postcentral gyrus, (E) caudal middle frontal cortex, and (F) banks of the superior temporal sulcus.

### Investigation of the effects of stimulation intensity and frequency

To determine the modulatory effects of stimulation amplitude and frequency on cortical activity, we compared different DBS settings at the discriminative nodes identified as part of the minimally discriminative functional network response profile. Firstly, unilateral stimulation had weaker modulatory effects than bilateral stimulation in the alpha and gamma frequency bands. Bilateral stimulation at 130 Hz and 1.5 mA was associated with undercompensated activity in non-responders and normalized or overcompensated activity in responders (Figure 5A). In contrast, unilateral stimulation was associated with undercompensated activity in non-responders and responders (Figure 5B).

**Figure 5.**
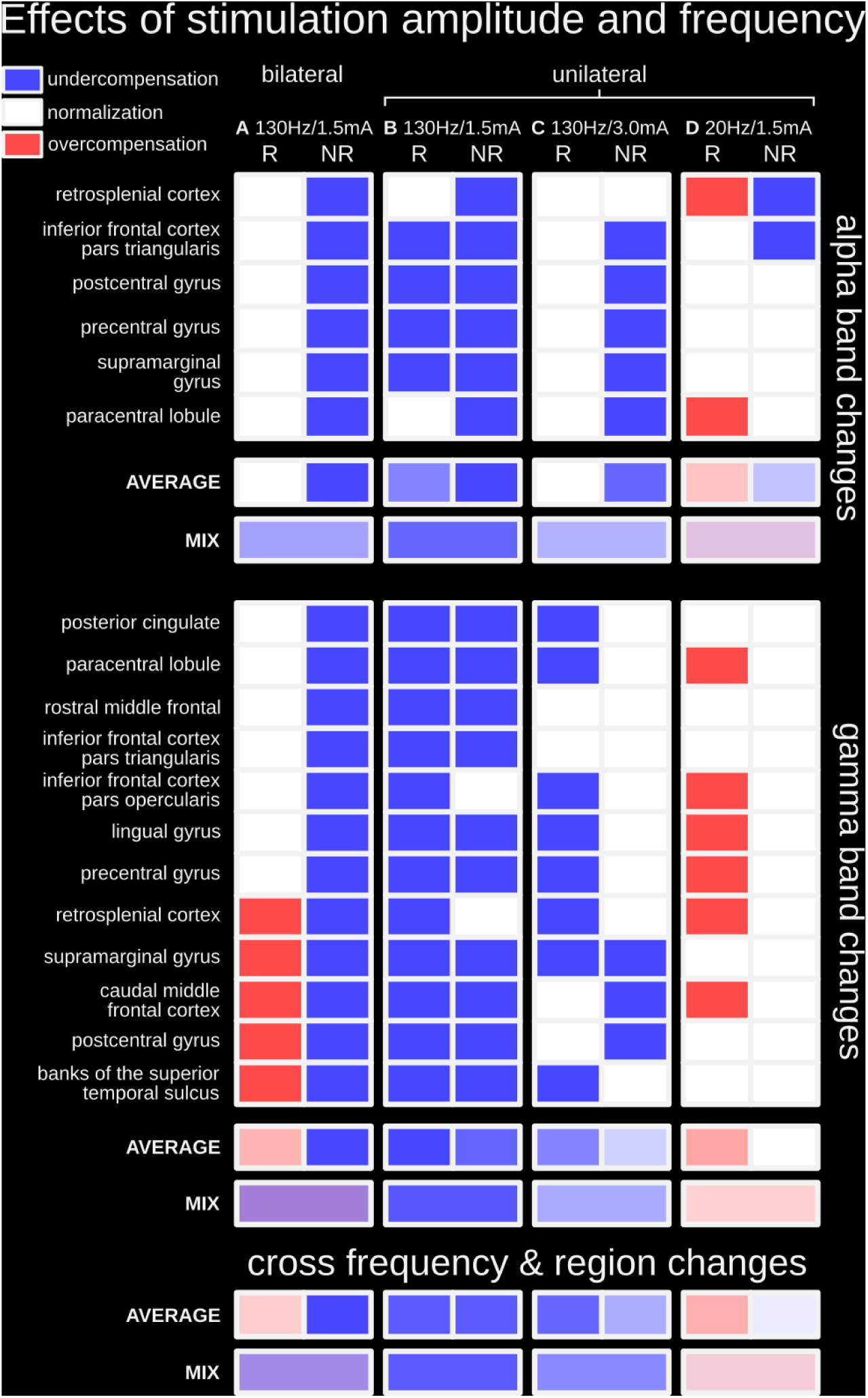
Investigation of the effects of stimulation amplitude and frequency. (A) Bilateral stimulation at 130 Hz/1.5 mA was associated with undercompensated activity in non-responders and normalized and/or overcompensated activity in responders; this column represents the statistical summary of Figures 2 – 4. (B) Unilateral stimulation at 130 Hz/1.5 mA was associated with generally undercompensated activity in both responders and non-responders in both alpha and gamma frequency bands; this condition is intended to serve as a new baseline to investigate the effects of stimulation amplitude and frequency. (C) Unilateral stimulation at 130 Hz using a higher amplitude of 3.0 mA (instead of 1.5 mA) led to strengthened modulatory effects (i.e., less undercompensated nodes). (D) Unilateral stimulation at 1.5 mA using a lower stimulation frequency of 20 Hz (instead of 130 Hz) led to even stronger modulatory effects (i.e., less undercompensated nodes and more overcompensated nodes). R = responders; NR = non-responders. Detailed statistical depictions are available in Supplementary Figures 2 & 3.

For stimulation amplitude, cortical modulatory effects at the discriminative nodes were more substantial with an increase in intensity. When unilateral stimulation at 130 Hz was increased from 1.5 to 3.0 mA, modulatory effects were strengthened overall (i.e., less undercompensated nodes; Figure 5C). In the alpha band, all discriminative nodes became normalized in responders, while effects remained generally undercompensated in non-responders. In the gamma band, while more nodes became normalized overall, this response was generally more often observed in non-responders. Statistical details for the cortical response to changes in stimulation amplitude are available in Supplementary Figures 2 & 3.

For stimulation frequency, cortical modulatory effects at the discriminative nodes were more substantial with a decrease in frequency. When unilateral stimulation at 1.5 mA was decreased from 130 to 20 Hz, modulatory effects were strengthened overall (i.e., less undercompensated nodes and more overcompensated nodes; Figure 5D). In the alpha and gamma bands, discriminative nodes became normalized or overcompensated in responders, whereas nodes became normalized or remained undercompensated in non-responders. The strengthening of modulatory effects was greater with titrating stimulation frequency than amplitude. Statistical details for the cortical response to changes in stimulation frequency are available in Supplementary Figures 2 & 3.

## Discussion

Currently, deep brain stimulation (DBS) parameter optimization in people with treatment resistant depression (TRD) is solely informed through subjective assessment tools (such as self-reports and clinician observations) and requires months of adjustment in order to determine optimal settings. Consequently, this strongly limits the number of DBS parameter combinations being trialed for TRD due to limited time and resources.

In this study, we lay the groundwork for this process to be augmented by the use of an objective measure, namely individualized electrophysiological feedback, to expedite the process. We identified electrophysiological profiles related to effective treatment responses which may be used to aid in DBS lead placement/contact selection and DBS pulse parameters (frequency/amplitude) in prospective applications of SCC-DBS. The use of electrophysiological feedback to determine electrode placement has long been considered a gold-standard for DBS in for movement disorders (16), and more recently, specific electrophysiological profiles (e.g. beta frequency peaks) have been shown to align with the clinically most effective contacts (17), suggesting that electrophysiological information can be used to guide stimulation programming (18, 19). The electrophysiological profiles identified herein may be employed for such a purpose, but in the context of TRD, to verify DBS lead placement and contact selection.

Additionally, we identified the electrophysiological profile change aligned with SCC-DBS amplitude (1.5 mA vs. 3 mA) and frequency (20 Hz vs. 130 Hz) changes. While low-frequency subthalamic stimulation (< 50 Hz) is ineffective/disadvantageous in movement disorders (20), this does not necessarily apply to psychiatric disorders such as TRD. As such, the potential search space for DBS parameter optimization is much larger, making DBS setting selection more complex in TRD. However, an understanding of the general neuromodulatory effects of SCC-DBS in TRD allows for an informed, electrophysiologically-guided search through the DBS pulse parameter space to quickly optimize treatment efficacy in a novel case.

The electrophysiological characteristics of the herein identified minimally discriminative functional network SCC-DBS response profile were characterized by a responder-specific modulation of pathological activity, differing dependent on DBS amplitude and frequency. Spatially, the network encompassed nodes of the default mode network (DMN), central executive network (CEN), and somatomotor network, as well as Broca’s area, lingual gyrus, and temporal areas. These regions are involved in emotional control and processing, motor control, and the interaction between speech, vision, and memory, which have all been implicated in depression (21–25). The spectral characteristics of these observations, namely amplified low-frequency and decreased high-frequency (gamma) oscillations, mirror previous reports on pathological changes in people with TRD (26).

### Modulated nodes in the default mode network

The DMN is, among other functions, involved in emotional processing (27) and has been implicated in depression, specifically TRD (28). Several cortical regions that are part of the DMN network were identified in our study as nodes of interest, including the posterior cingulate cortex (PCC), retrosplenial cortex (RSC) (29), and supramarginal gyrus (SG) (30). Triggered by emotionally linked cues, interactions between structures within the DMN have been observed during the processing of emotional experiences (31) and contextual learning in rodents (32). In addition, the DMN plays a crucial role in memory formation and emotion-memory linkage. During resting periods, the network processes memory-related information in self-referential tasks such as introspection, worry, rumination (33), and other memory-related tasks (spatial and episodic) (34, 35). The RSC, in particular, is a node in the DMN that is anatomically well-situated to fuse emotions and memory (36) and has been implicated in fear conditioning (37) and contextual learning (32) in rodents. Furthermore, increased RSC low-frequency (4-12 Hz) and gamma activities have been associated with mnemonic functions (37). Complementary evidence from human studies suggests RSC blood flow (38) and BOLD activity increase during emotional processing (39). Increased RSC BOLD activity has also been linked to modulations in RSC alpha power (40). Generally, RSC and PCC are commonly activated in emotion processing(36), as is the SG. The SG has also been shown to be activated in tasks related to emotion identification (41).

Furthermore, pathological changes within the DMN have been linked to several psychiatric/ neurological disorders such as Alzheimer’s disease (42), major depression (24), obsessive-compulsive disorder (43), social phobia (44), and schizophrenia (45). While metabolic activity in the RSC/PCC is decreased in Alzheimer’s disease (42), it is pathologically increased in people with depression (46). Additionally, increased depressive symptom severity has been associated with increased left RSC/PCC volume (47). Similarly, cortical thickening has been observed in the SG of people with major depressive disorder (25). Overall, the aforementioned studies support the potential importance of modulating certain disease-relevant nodes of the DMN uncovered by this work, namely, responder-specific normalizations of pathologically increased alpha power (RSC and SG) and decreased gamma power (PCC).

### Modulated nodes in the central executive network

The CEN is involved in attention control, working memory, and decision-making (48). It primarily consists of the dorsolateral prefrontal cortex (dlPFC) and posterior parietal cortex (PPC) (49). Although activity in the CEN is closely connected to the DMN, the two networks display anticorrelated behavior (50). Gamma band activity in the CEN, particularly the dlPFC, has been connected with working memory performance (51). Likewise, decreased gamma activity in the dlPFC has also been associated with reduced cognitive control and blunted reward learning in people with depression (52). Reduced glucose metabolic rates have also been observed in the left dlPFC of people with depression symptoms (21). These studies reflect our observation of pathologically decreased gamma power in the left rostral middle frontal cortex (RMFC), which contains the left dlPFC. In particular, we found a responder-specific normalization of pathologically decreased gamma band power in the left RMFC/dlPFC region with SCC-DBS. Similar results have been reported when applying transcranial magnetic stimulation (TMS) to the left dlPFC (53). As such, the left dlPFC is a common target for TMS in depression treatment (54), likely due to its role in dopamine release (55) and its connections to several cortical and subcortical regions implicated in depression (56).

### Modulated nodes of other networks

In responders, SCC-DBS was also associated with the modulation of other brain regions and networks that are functionally and structurally connected to the DMN and CEN. One such network that was pathologically implicated in our study was the somatomotor network (pre- and postcentral gyri) and surrounding areas (paracentral lobule). Pathological alterations have previously been shown throughout this network in people with depression (57), such as increases in white matter hyperintensities (58). These observations have often been linked to psychomotor retardation (58), a common symptom of depression (59). Our study identified responder-specific normalization of pathologically increased alpha power (pre- and postcentral gyri and paracentral lobule) and decreased gamma power (paracentral lobule). Acute modulatory effects in these regions are likely mediated by trans-synaptic connectivity to SCC via the thalamus (60, 61).

We also observed responder-specific normalization of increased alpha power (pars triangularis) and decreased gamma power (pars triangularis and opercularis) within the inferior frontal cortex. Disease-related changes in altered anatomy have been reported in the pars opercularis (25) and triangularis (62) (forming Broca’s area) in people with depression. Moreover, in people with depression, reduced oscillatory beta-band power was measured in the vicinity of Broca’s area, relating to the error rates in phonological tasks (63). Anatomically, these nodes within Broca’s area are connected to the amygdala and are thought to play a role in a top-down control mechanism in worrying (64). The amygdala, in turn, has reciprocal connections to the SCC, indirectly linking the nodes of Broca’s area to the target of SCC-DBS.

We also identified responder-specific normalization of pathologically decreased gamma power in the lingual gyrus. This region is thought to play an important role in abnormal visual processing in people with depression (22). Larger gray matter volumes of the lingual gyrus have been associated with better performance in neuropsychological tests (22). Functionally, activations of the lingual gyrus have been connected to activations of the amygdala (65).

Lastly, we observed responder-specific overcompensation of pathologically decreased gamma power in the left superior temporal sulcus and left caudal middle frontal gyrus. Changes to the superior temporal sulcus may be associated with sleep quality in depression (66).

### Limitations

This study applied SCC-DBS using a unified parameter set to identify a minimally discriminative functional network response profile. As such, patient-specific parameters determined clinically were not used in the study. While stimulation frequency and pulse width were used per conventional clinical settings, bipolar stimulation was employed to limit stimulation artifacts during MEG recordings. In addition, a unified stimulation amplitude was used to identify an electrophysiological profile that may be used to optimize treatment delivery in prospective contexts. This would otherwise not be possible if using heterogeneous stimulation settings. While this approach cannot be used to calibrate DBS amplitude per se, it provides a viable avenue for DBS lead position verification and contact selection. The analyses led to identifying a discriminative network profile derived from 3-minute MEG recordings, which may be informative of responder status. However, the patient sample size was limited, and MEG recordings were acquired with *a priori* knowledge of responder status. As such, the potential clinical utility of the identified discriminative network profile would need to be established by obtaining MEG scans at an early stage in therapy initiation to guide DBS programming. Additionally, although our results indicate that low-frequency SCC-DBS may have the strongest counter pathological effects, clinical applicability is yet to be confirmed. Furthermore, it is necessary to consider that TRD is a highly heterogeneous disorder with variable symptomatology and associated electrophysiological profiles, whereas our analysis is limited to a binary discretization of responder status due to the limited data in this retrospective study. Moreover, our interpretations of the presented findings are limited to the identified areas of interest and their reported relevance in TRD but do not provide an exhaustive overview of all potentially involved cortical regions, their functions, or pathological alterations in TRD.

## Conclusion

This study is among the first to use MEG to examine whole-brain and region-specific cortical responses to SCC-DBS in people with TRD. The application of SCC-DBS at unified parameter settings led to the identification of a minimally discriminative network profile that could differentiate responders from non-responders characterized by modulations of increased alpha and decreased gamma power in nodes of the DMN, CEN, and somatomotor network, as well as Broca’s area and lingual gyrus – regions that have been implicated in behavioral functions and impairments associated with depression. The identified discriminative network profile may represent a functional readout that can be used for optimizing SCC-DBS therapy via improved candidate selection, surgical targeting, and DBS setting selection in prospective contexts. Furthermore, our mechanistic interrogations of stimulation settings revealed that increasing stimulation amplitude strengthened modulatory network effects, while lowering stimulation frequency had even more profound neuromodulatory effects.

## Competing interests

A.M.L. is a consultant to Abbott, Boston Scientific, Medtronic, and Functional Neuromodulation. L.M. has received honoraria and travel funds from Medtronic (unrelated to this work). All other authors declare no competing interests.

Supplementary material is available online.

