## Supplementary Material for "Cortical network mechanisms in subcallosal cingulate deep brain stimulation for depression"

### Supplementary Methods

#### Surgical procedure

Bilateral SCC-DBS implantation was performed at the University Health Network-Toronto Western Hospital (Toronto, ON, Canada). Electrode coordinates were based on 3T structural T1- and T2-weighted magnetic resonance imaging (MRI). Routine microelectrode recording procedures preceded the insertion of electrodes (4-contact lead, Abbott (formerly St. Jude Medical), Illinois, USA) in the SCC under local anesthesia. As per the standard of care, all patients underwent a postoperative MRI for electrode localization. All electrode locations were confirmed to be within normal limits in all subjects, and no abnormalities were noted.

#### Postoperative lead localization

DBS electrode placement was estimated on the basis of patient-specific structural imaging using Lead-DBS [1], as previously described [2–4]. Briefly, each patient's pre-operative T1-weighted MR image was rigidly coregistered with a corresponding post-operative structural image (either MRI or CT). Post-operative brain shift was corrected for as needed using an additional affine transformation. The coregistered images were then normalized to MNI space (MNI152 NLIN 2009b, Montreal Neurological Institute) [5, 6] using the nonlinear 'effective low variance' ANTs SyN algorithm with subcortical refinement (http://stnava.github.io/ANTs/ [7]). Finally, DBS leads were localized on the post-operative images by way of semiautomated trajectory reconstruction with additional manual refinement, before being transformed to MNI space via the nonlinear warp field generated earlier.

#### MEG recordings & paradigm

Data were recorded using a 306 channel MEG unit (Elekta Neuromag TRIUX$^{TM}$, Helsinki, Finland) at a frequency sampling rate of 1,000 Hz with an online bandpass filter between 0.1 and 330 Hz.

Recording occurred while participants were supine with their eyes closed at rest. To determine the effect of SCC-DBS on cortical activity, patients underwent three-minute recordings with DBS OFF and DBS ON. Bipolar DBS settings were used to avoid sensor saturation. The remaining SCC-DBS parameters were configured as follows: 130 Hz frequency, 90 μs pulse width, 1.5 mA amplitude, and central contacts (1-2+) configuration. Artifacts from nearby sources in the MEG data were removed using a temporal signal space separation (tSSS) algorithm (Elekta Neuromag MaxFilter software version 2.2.12, Elekta, Helsinki, Finland) [8–10] with the default 10 second time window and subspace correlation limit of 0.980.

#### Data pre-processing

The raw data were filtered using a finite impulse response filter. This filter notched line noise at 60 Hz and its harmonic at 120 Hz. Spectral content above 120 Hz was also removed to (1) avoid subsequent harmonics of line noise, (2) avoid DBS artifacts at 130 Hz, and (3) support downsampling the signal to 240 Hz to increase data processing speeds.

#### Source reconstruction

Before extracting features from the data, recordings were transformed from sensor to source space. Each subject's source space was also transformed into a common source space, enabling comparison across subjects. Head models were computed and coregistered with the recorded data as the first step. For TRD patients, individual MRI scans were available. Cortical source space and boundary element models (BEM) models were extracted using FreeSurfer (v7.2.0, [11]). The validity of this extraction process was visually confirmed. For healthy controls, MRI scans were unavailable. Therefore, fs-average (an experimentally derived standard head model provided within FreeSurfer) was used as a head model. Recordings were coregistered (using MNE [12]) to the extracted models (TRD subjects) or fs-average (for HC subjects). After visually verifying the success of the coregistration for every recording, an inverse operator was calculated and subsequently applied to transform the data from sensor to source space. All data were then morphed into the source space of fs-average. To conserve statistical power and to maintain reasonable processing speed, all channels within individual cortical regions outlined by the Desikan-Killiany atlas were averaged to represent their respective regions (Desikan-Killiany atlas [13], see Supplementary Figure 1). Afterwards, the data were further sub-epoched into 15 s and then 30 s to investigate the impact of different epoching on the evaluation results. Finally, relevant data were transformed [14] and exported into Blender (v3.1.2; [15]) for visualization.

#### Power analyses

Spectral power estimates were extracted from common source space transformed data, the delta (2-4 Hz), theta (4-8 Hz), alpha (8-12 Hz), beta (12-32 Hz), and gamma bands (32-116 Hz) were explored. Although activity within these frequency bands is commonly associated with specific neurological processes, peaks within these bands do not commonly span the entire respective frequency band. We leveraged this characteristic to reduce the signal to noise ratio in our data by having a sub-band represent peak activity for the overall frequency band. To this end, each frequency band was subdivided into a several of different sub-bands. Delta activity was divided into $delta_1$ (2-3 Hz), $delta_2$ (2.5-3.5 Hz), and $delta_3$ (3-4 Hz) sub-bands. Theta was divided into $theta_1$ (4-6 Hz), $theta_2$ (5-7 Hz), and $theta_3$ (6-8 Hz) sub-bands. Alpha was divided into $alpha_1$ (8-10 Hz), $alpha_2$ (9-11 Hz), and $alpha_3$ (10-12Hz) sub-bands. Beta was divided into $beta_1$ (12-16 Hz), $beta_2$ (14-18 Hz), $beta_3$ (16-20 Hz), $beta_4$ (18-22 Hz), $beta_5$ (20-24 Hz), $beta_6$ (22-26 Hz), $beta_7$ (24-28 Hz), $beta_8$ (26-30 Hz), and $beta_9$ (28-32 Hz). Gamma activity was divided into 8 Hz wide sub-bands, starting with $gamma_1$ (32-40 Hz) until $gamma_{20}$ (108-116 Hz) sub-bands. Smaller step sizes were chosen for low-frequency bands compared to higher frequency bands as a 1 Hz change in a low-frequency band (e.g., theta) is relatively much wider than in a high-frequency band (e.g., gamma). Power in each sub-band was estimated as the ratio of the absolute sum of all activity within a sub-band, divided by the absolute sum of all activity in the broad-band (2-120 Hz). Normalizing power estimates towards the overall power compensated for different recording sensitivities across channels within and across patients. Finally, to derive power estimates for non-sub-bands (e.g., theta), the sub-band with the highest power (e.g., $theta_2$) was chosen as the power estimate of this frequency band. Discriminative nodes were statistically identified as described in the main text of the manuscript.

### Supplementary Figures

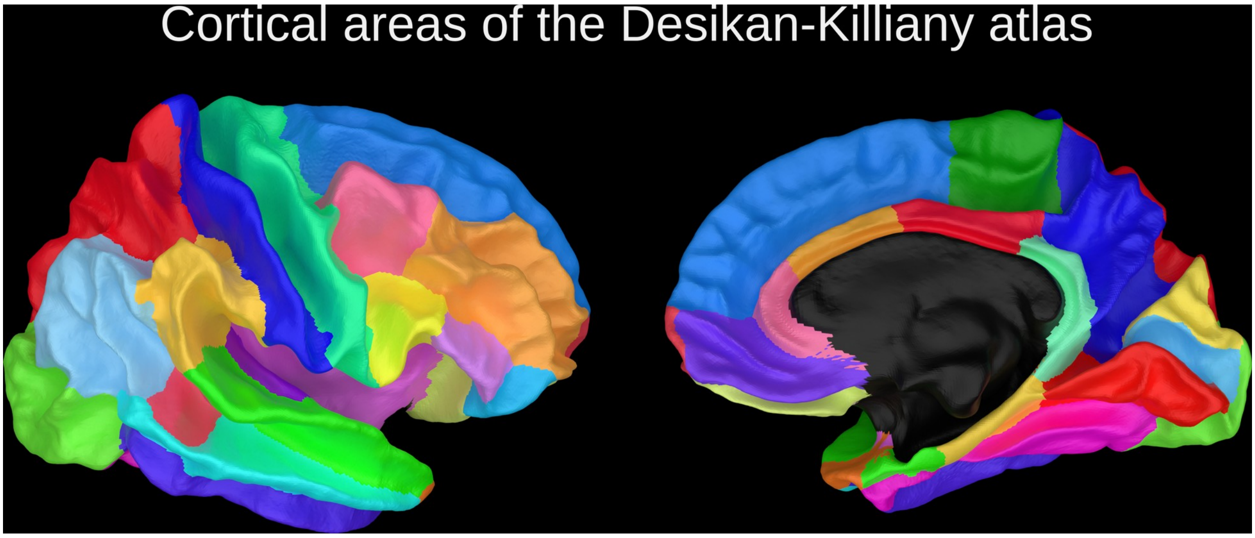

**Supplementary Figure 1 – Cortical areas of the Desikan-Killiany atlas.** Individual regions are separated by color (black region not investigated).

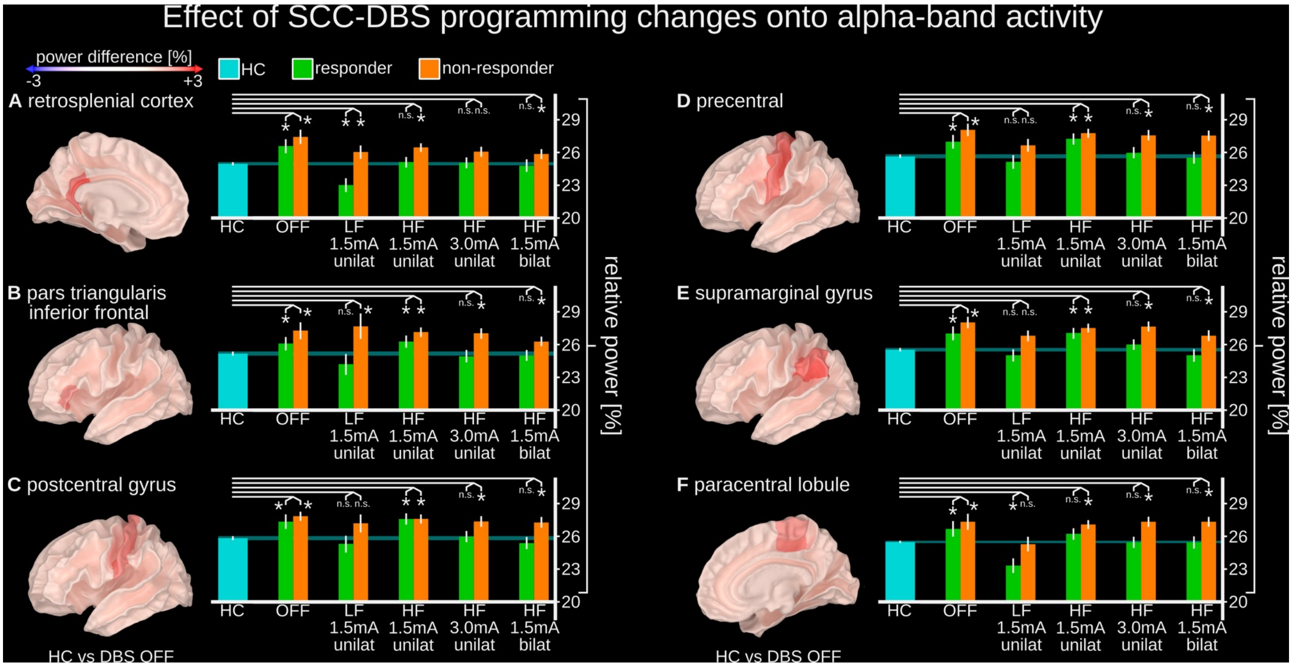

**Supplementary Figure 2 – Effect of SCC-DBS parameter changes on alpha-band activity.** The figure depicts detailed statistical results on a region-by-region basis; whereas the effects are summarized within Figure 5 of the main text.

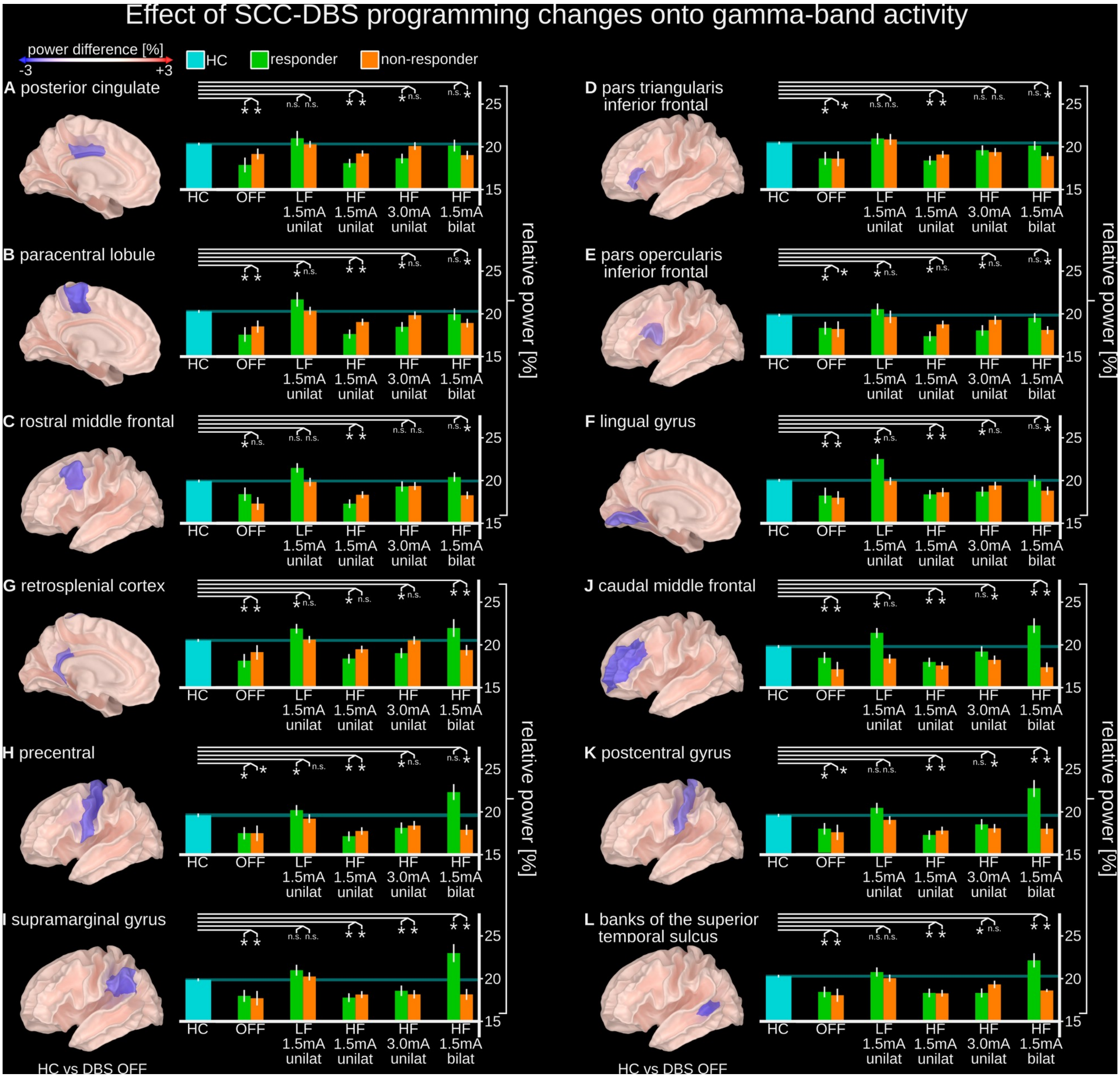

**Supplementary Figure 3 – Effect of SCC-DBS parameter changes on gamma-band activity. The figure depicts detailed statistical results on a region-by-region basis; whereas the effects are summarized within Figure 5 of the main text.**

### Supplementary Table 1 – Additional clinical information

| Subject | # Suicide attempts | # Family members history of MDD | Past therapies | | |
|---|---|---|---|---|---|
| | | | Psychotherapy | ECT | rTMS |
| 1 | 0-3 | 0-3 | yes | yes | yes |
| 2 | 0-3 | 0-3 | yes | yes | no |
| 3 | 0-3 | 0-3 | yes | yes | yes |
| 4 | 0-3 | 0-3 | yes | no | yes |
| 5 | 0-3 | 0-3 | yes | yes | no |
| 6 | 0-3 | 4-6 | yes | no | no |
| 7 | 0-3 | 4-6 | yes | no | yes |
| 8 | 0-3 | 0-3 | yes | yes | no |
| 9 | 0-3 | 0-3 | yes | yes | no |
| 10 | 0-3 | 0-3 | yes | yes | no |
| 11 | 0-3 | 0-3 | yes | no | yes |
| 12 | 0-3 | 0-3 | yes | yes | yes |
| 13 | 0-3 | 0-3 | yes | yes | no |
| 14 | * | * | yes | * | * |
| 15 | 4-6 | 0-3 | yes | yes | yes |

**DBS, deep brain stimulation; ECT, electroconvulsive therapy; HAM-D17, 17-item Hamilton Depression Rating Scale; MDD, major depressive disorder; rTMS, repetitive transcranial magnetic stimulation; SCC, subcallosal cingulate gyrus; SNRI, serotonin and norepinephrine reuptake inhibitor; SSRI, selective serotonin reuptake inhibitor; TRD, treatment-resistant depression.**

### Supplementary Table 2 – Additional statistical information

| Retrosplenial – left hemisphere – alpha band | | | | | | | | | |
|---|---|---|---|---|---|---|---|---|---|
| **Compared factor phenotypes** | | | epoch size [s] | values | | variance | | significance | |
| phenotype A | phenotype B | | - | phen. A | phen. B | phen. A | phen. B | phen. A | intercept |
| resp_dbs_off | resp_dbs_bilat | | 15 | 0.266 | 0.248 | 0.011 | 0.004 | 1.02E-04 | 7.63E-128 |
| resp_dbs_off | resp_dbs_bilat | | 30 | 0.265 | 0.247 | 0.011 | 0.006 | 3.29E-03 | 7.85E-134 |
| nresp_dbs_off | nresp_dbs_bilat | | 15 | 0.273 | 0.257 | 0.010 | 0.004 | 3.11E-05 | 3.59E-149 |
| nresp_dbs_off | nresp_dbs_bilat | | 30 | 0.272 | 0.256 | 0.011 | 0.005 | 4.63E-04 | 3.38E-140 |
| hc | resp_dbs_bilat | | 15 | 0.250 | 0.248 | 0.001 | 0.004 | 5.24E-01 | 0.00E+00 |
| hc | resp_dbs_bilat | | 30 | 0.248 | 0.247 | 0.002 | 0.005 | 7.82E-01 | 3.69E-304 |
| hc | nresp_dbs_bilat | | 15 | 0.250 | 0.260 | 0.001 | 0.004 | 8.31E-03 | 0.00E+00 |
| hc | nresp_dbs_bilat | | 30 | 0.248 | 0.259 | 0.002 | 0.005 | 2.95E-02 | 0.00E+00 |
| resp | nresp_dbs_bilat | | 15 | 0.260 | 0.248 | 0.005 | 0.006 | 6.57E-02 | 9.11E-108 |
| resp | nresp_dbs_bilat | | 30 | 0.259 | 0.247 | 0.006 | 0.009 | 1.85E-01 | 2.88E-57 |
| hc | resp_dbs_off | | 15 | 0.250 | 0.265 | 0.001 | 0.003 | 1.03E-05 | 0.00E+00 |
| hc | resp_dbs_off | | 30 | 0.248 | 0.266 | 0.002 | 0.004 | 1.23E-04 | 0.00E+00 |
| hc | nresp_dbs_off | | 15 | 0.250 | 0.275 | 0.001 | 0.003 | 1.55E-12 | 0.00E+00 |
| hc | nresp_dbs_off | | 30 | 0.248 | 0.275 | 0.002 | 0.004 | 4.12E-09 | 0.00E+00 |
| resp | nresp_dbs_off | | 15 | 0.275 | 0.265 | 0.003 | 0.005 | 6.33E-02 | 8.60E-145 |
| resp | nresp_dbs_off | | 30 | 0.275 | 0.266 | 0.005 | 0.007 | 1.72E-01 | 5.68E-78 |
| Triangularis – left hemisphere – alpha band | | | | | | | | | |
| Compared factor phenotypes | | | epoch size [s] | values | | variance | | significance | |
| phenotype A | phenotype B | | - | phen. A | phen. B | phen. A | phen. B | phen. A | intercept |
| resp_dbs_off | resp_dbs_bilat | | 15 | 0.261 | 0.251 | 0.008 | 0.004 | 1.91E-02 | 1.81E-263 |
| resp_dbs_off | resp_dbs_bilat | | 30 | 0.260 | 0.248 | 0.008 | 0.005 | 1.51E-02 | 9.93E-253 |
| nresp_dbs_off | nresp_dbs_bilat | | 15 | 0.273 | 0.265 | 0.009 | 0.004 | 4.70E-02 | 5.66E-196 |
| nresp_dbs_off | nresp_dbs_bilat | | 30 | 0.269 | 0.262 | 0.009 | 0.004 | 6.36E-02 | 5.13E-192 |
| hc | resp_dbs_bilat | | 15 | 0.253 | 0.252 | 0.001 | 0.004 | 7.63E-01 | 0.00E+00 |
| hc | resp_dbs_bilat | | 30 | 0.250 | 0.248 | 0.002 | 0.005 | 6.71E-01 | 1.04E-297 |
| hc | nresp_dbs_bilat | | 15 | 0.253 | 0.263 | 0.001 | 0.004 | 1.29E-02 | 0.00E+00 |
| hc | nresp_dbs_bilat | | 30 | 0.250 | 0.261 | 0.002 | 0.005 | 4.55E-02 | 8.84E-294 |
| resp | nresp_dbs_bilat | | 15 | 0.263 | 0.252 | 0.004 | 0.005 | 2.58E-02 | 1.10E-123 |
| resp | nresp_dbs_bilat | | 30 | 0.261 | 0.248 | 0.005 | 0.007 | 5.57E-02 | 1.23E-66 |
| hc | resp_dbs_off | | 15 | 0.253 | 0.261 | 0.001 | 0.004 | 4.21E-02 | 0.00E+00 |
| hc | resp_dbs_off | | 30 | 0.250 | 0.260 | 0.002 | 0.005 | 4.23E-02 | 5.64E-299 |
| hc | nresp_dbs_off | | 15 | 0.253 | 0.275 | 0.001 | 0.004 | 1.24E-08 | 0.00E+00 |
| hc | nresp_dbs_off | | 30 | 0.250 | 0.271 | 0.002 | 0.005 | 1.58E-05 | 3.63E-304 |
| resp | nresp_dbs_off | | 15 | 0.275 | 0.261 | 0.003 | 0.005 | 6.02E-03 | 7.96E-149 |
| resp | nresp_dbs_off | | 30 | 0.271 | 0.260 | 0.004 | 0.006 | 7.32E-02 | 4.56E-83 |
| Postcentral – left hemisphere – alpha band | | | | | | | | | |
| Compared factor phenotypes | | | epoch size [s] | values | | variance | | significance | |
| phenotype A | phenotype B | | - | phen. A | phen. B | phen. A | phen. B | phen. A | intercept |
| resp_dbs_off | resp_dbs_bilat | | 15 | 0.274 | 0.254 | 0.011 | 0.004 | 3.44E-06 | 1.26E-135 |
| resp_dbs_off | resp_dbs_bilat | | 30 | 0.271 | 0.253 | 0.011 | 0.005 | 5.89E-04 | 1.86E-146 |
| nresp_dbs_off | nresp_dbs_bilat | | 15 | 0.278 | 0.274 | 0.007 | 0.004 | 3.82E-01 | 0.00E+00 |
| nresp_dbs_off | nresp_dbs_bilat | | 30 | 0.277 | 0.274 | 0.008 | 0.005 | 5.84E-01 | 1.04E-260 |
| hc | resp_dbs_bilat | | 15 | 0.259 | 0.254 | 0.002 | 0.004 | 2.86E-01 | 0.00E+00 |
| hc | resp_dbs_bilat | | 30 | 0.257 | 0.253 | 0.002 | 0.006 | 4.30E-01 | 7.72E-290 |
| hc | nresp_dbs_bilat | | 15 | 0.259 | 0.271 | 0.002 | 0.004 | 4.69E-03 | 0.00E+00 |
| hc | nresp_dbs_bilat | | 30 | 0.257 | 0.272 | 0.002 | 0.006 | 1.30E-02 | 5.46E-289 |
| resp | nresp_dbs_bilat | | 15 | 0.271 | 0.254 | 0.004 | 0.006 | 3.43E-03 | 4.02E-115 |
| resp | nresp_dbs_bilat | | 30 | 0.272 | 0.253 | 0.006 | 0.008 | 1.91E-02 | 1.45E-62 |
| hc | resp_dbs_off | | 15 | 0.259 | 0.275 | 0.002 | 0.004 | 2.11E-04 | 0.00E+00 |
| hc | resp_dbs_off | | 30 | 0.257 | 0.271 | 0.002 | 0.005 | 1.11E-02 | 8.36E-293 |

| | | | | | | | | | |
|---|---|---|---|---|---|---|---|---|---|
| hc | nresp_dbs_off | | 15 | 0.259 | 0.278 | 0.002 | 0.004 | 1.34E-06 | 0.00E+00 |
| hc | nresp_dbs_off | | 30 | 0.257 | 0.278 | 0.002 | 0.005 | 3.92E-05 | 1.14E-300 |
| resp | nresp_dbs_off | | 15 | 0.278 | 0.275 | 0.003 | 0.005 | 4.37E-01 | 1.72E-147 |
| resp | nresp_dbs_off | | 30 | 0.278 | 0.271 | 0.004 | 0.006 | 2.63E-01 | 2.61E-81 |
| Precentral – left hemisphere – alpha band | | | | | | | | | |
| Compared factor phenotypes | | | epoch size [s] | values | | variance | | significance | |
| phenotype A | phenotype B | | - | phen. A | phen. B | phen. A | phen. B | phen. A | intercept |
| resp_dbs_off | resp_dbs_bilat | | 15 | 0.271 | 0.255 | 0.010 | 0.004 | 2.50E-04 | 2.13E-160 |
| resp_dbs_off | resp_dbs_bilat | | 30 | 0.268 | 0.255 | 0.010 | 0.005 | 1.74E-02 | 1.47E-165 |
| nresp_dbs_off | nresp_dbs_bilat | | 15 | 0.280 | 0.277 | 0.007 | 0.004 | 5.13E-01 | 0.00E+00 |
| nresp_dbs_off | nresp_dbs_bilat | | 30 | 0.279 | 0.275 | 0.008 | 0.005 | 3.69E-01 | 4.39E-275 |
| hc | resp_dbs_bilat | | 15 | 0.257 | 0.255 | 0.001 | 0.004 | 6.00E-01 | 0.00E+00 |
| hc | resp_dbs_bilat | | 30 | 0.255 | 0.255 | 0.002 | 0.005 | 9.85E-01 | 9.77E-299 |
| hc | nresp_dbs_bilat | | 15 | 0.257 | 0.277 | 0.001 | 0.004 | 3.99E-06 | 0.00E+00 |
| hc | nresp_dbs_bilat | | 30 | 0.255 | 0.273 | 0.002 | 0.005 | 8.29E-04 | 2.64E-298 |
| resp | nresp_dbs_bilat | | 15 | 0.277 | 0.255 | 0.004 | 0.006 | 2.89E-04 | 9.06E-117 |
| resp | nresp_dbs_bilat | | 30 | 0.273 | 0.255 | 0.005 | 0.008 | 2.26E-02 | 3.67E-63 |
| hc | resp_dbs_off | | 15 | 0.257 | 0.271 | 0.001 | 0.004 | 3.01E-04 | 0.00E+00 |
| hc | resp_dbs_off | | 30 | 0.255 | 0.268 | 0.002 | 0.005 | 1.26E-02 | 1.00E-303 |
| hc | nresp_dbs_off | | 15 | 0.257 | 0.281 | 0.001 | 0.004 | 3.07E-10 | 0.00E+00 |
| hc | nresp_dbs_off | | 30 | 0.255 | 0.280 | 0.002 | 0.005 | 1.14E-07 | 0.00E+00 |
| resp | nresp_dbs_off | | 15 | 0.281 | 0.271 | 0.003 | 0.005 | 4.15E-02 | 6.73E-154 |
| resp | nresp_dbs_off | | 30 | 0.280 | 0.268 | 0.004 | 0.006 | 3.22E-02 | 1.83E-84 |
| Supramarginal – left hemisphere – alpha band | | | | | | | | | |
| Compared factor phenotypes | | | epoch size [s] | values | | variance | | significance | |
| phenotype A | phenotype B | | - | phen. A | phen. B | phen. A | phen. B | phen. A | intercept |
| resp_dbs_off | resp_dbs_bilat | | 15 | 0.270 | 0.249 | 0.011 | 0.004 | 4.69E-07 | 1.55E-131 |
| resp_dbs_off | resp_dbs_bilat | | 30 | 0.269 | 0.251 | 0.011 | 0.006 | 1.15E-03 | 2.06E-136 |
| nresp_dbs_off | nresp_dbs_bilat | | 15 | 0.281 | 0.270 | 0.010 | 0.004 | 1.14E-02 | 3.69E-189 |
| nresp_dbs_off | nresp_dbs_bilat | | 30 | 0.277 | 0.268 | 0.009 | 0.005 | 8.18E-02 | 9.07E-190 |
| hc | resp_dbs_bilat | | 15 | 0.256 | 0.249 | 0.001 | 0.004 | 1.18E-01 | 0.00E+00 |
| hc | resp_dbs_bilat | | 30 | 0.254 | 0.251 | 0.002 | 0.005 | 5.10E-01 | 1.04E-296 |
| hc | nresp_dbs_bilat | | 15 | 0.256 | 0.268 | 0.001 | 0.004 | 2.59E-03 | 0.00E+00 |
| hc | nresp_dbs_bilat | | 30 | 0.254 | 0.266 | 0.002 | 0.005 | 2.78E-02 | 1.03E-296 |
| resp | nresp_dbs_bilat | | 15 | 0.268 | 0.249 | 0.005 | 0.007 | 4.34E-03 | 1.26E-107 |
| resp | nresp_dbs_bilat | | 30 | 0.266 | 0.251 | 0.006 | 0.009 | 8.20E-02 | 3.04E-58 |
| hc | resp_dbs_off | | 15 | 0.256 | 0.271 | 0.001 | 0.004 | 1.56E-04 | 0.00E+00 |
| hc | resp_dbs_off | | 30 | 0.254 | 0.269 | 0.002 | 0.005 | 2.74E-03 | 2.16E-304 |
| hc | nresp_dbs_off | | 15 | 0.256 | 0.282 | 0.001 | 0.004 | 3.79E-12 | 0.00E+00 |
| hc | nresp_dbs_off | | 30 | 0.254 | 0.279 | 0.002 | 0.005 | 1.50E-07 | 0.00E+00 |
| resp | nresp_dbs_off | | 15 | 0.282 | 0.271 | 0.003 | 0.005 | 2.04E-02 | 1.32E-149 |
| resp | nresp_dbs_off | | 30 | 0.279 | 0.269 | 0.004 | 0.006 | 1.09E-01 | 6.90E-82 |
| Paracentral – right hemisphere – alpha band | | | | | | | | | |
| Compared factor phenotypes | | | epoch size [s] | values | | variance | | significance | |
| phenotype A | phenotype B | | - | phen. A | phen. B | phen. A | phen. B | phen. A | intercept |
| resp_dbs_off | resp_dbs_bilat | | 15 | 0.266 | 0.254 | 0.012 | 0.005 | 9.35E-03 | 1.05E-105 |
| resp_dbs_off | resp_dbs_bilat | | 30 | 0.267 | 0.254 | 0.011 | 0.006 | 4.00E-02 | 4.84E-126 |
| nresp_dbs_off | nresp_dbs_bilat | | 15 | 0.271 | 0.268 | 0.011 | 0.004 | 3.86E-01 | 4.21E-133 |
| nresp_dbs_off | nresp_dbs_bilat | | 30 | 0.270 | 0.266 | 0.011 | 0.004 | 3.16E-01 | 4.59E-137 |
| hc | resp_dbs_bilat | | 15 | 0.255 | 0.254 | 0.001 | 0.004 | 7.47E-01 | 0.00E+00 |
| hc | resp_dbs_bilat | | 30 | 0.254 | 0.254 | 0.002 | 0.005 | 9.69E-01 | 1.84E-299 |
| hc | nresp_dbs_bilat | | 15 | 0.255 | 0.268 | 0.001 | 0.004 | 1.38E-03 | 0.00E+00 |
| hc | nresp_dbs_bilat | | 30 | 0.254 | 0.267 | 0.002 | 0.005 | 1.25E-02 | 2.05E-303 |
| resp | nresp_dbs_bilat | | 15 | 0.268 | 0.254 | 0.004 | 0.006 | 1.87E-02 | 7.36E-115 |
| resp | nresp_dbs_bilat | | 30 | 0.267 | 0.254 | 0.005 | 0.007 | 8.54E-02 | 1.30E-63 |
| hc | resp_dbs_off | | 15 | 0.255 | 0.266 | 0.001 | 0.004 | 7.06E-03 | 0.00E+00 |

| | | | | | | | | | |
|---|---|---|---|---|---|---|---|---|---|
| hc | resp_dbs_off | | 30 | 0.254 | 0.267 | 0.002 | 0.005 | 1.03E-02 | 2.04E-300 |
| hc | nresp_dbs_off | | 15 | 0.255 | 0.275 | 0.001 | 0.004 | 4.83E-07 | 0.00E+00 |
| hc | nresp_dbs_off | | 30 | 0.254 | 0.273 | 0.002 | 0.005 | 9.18E-05 | 7.61E-304 |
| resp | nresp_dbs_off | | 15 | 0.275 | 0.266 | 0.004 | 0.006 | 1.37E-01 | 7.89E-133 |
| resp | nresp_dbs_off | | 30 | 0.273 | 0.267 | 0.005 | 0.008 | 4.47E-01 | 7.97E-71 |
| bankssts – left hemisphere – gamma band | | | | | | | | | |
| Compared factor phenotypes | | | epoch size [s] | values | | variance | | significance | |
| phenotype A | phenotype B | | - | phen. A | phen. B | phen. A | phen. B | phen. A | intercept |
| resp_dbs_off | resp_dbs_bilat | | 15 | 0.188 | 0.223 | 0.019 | 0.006 | 3.08E-09 | 3.33E-22 |
| resp_dbs_off | resp_dbs_bilat | | 30 | 0.186 | 0.220 | 0.020 | 0.008 | 1.17E-05 | 1.66E-21 |
| nresp_dbs_off | nresp_dbs_bilat | | 15 | 0.180 | 0.188 | 0.015 | 0.003 | 6.30E-03 | 4.67E-33 |
| nresp_dbs_off | nresp_dbs_bilat | | 30 | 0.179 | 0.185 | 0.015 | 0.004 | 1.62E-01 | 3.23E-33 |
| hc | resp_dbs_bilat | | 15 | 0.203 | 0.222 | 0.002 | 0.005 | 2.69E-04 | 0.00E+00 |
| hc | resp_dbs_bilat | | 30 | 0.202 | 0.220 | 0.002 | 0.007 | 6.99E-03 | 5.81E-231 |
| hc | nresp_dbs_bilat | | 15 | 0.203 | 0.187 | 0.001 | 0.004 | 9.98E-05 | 0.00E+00 |
| hc | nresp_dbs_bilat | | 30 | 0.202 | 0.184 | 0.002 | 0.006 | 1.85E-03 | 1.68E-255 |
| resp | nresp_dbs_bilat | | 15 | 0.187 | 0.222 | 0.007 | 0.010 | 1.09E-03 | 3.07E-57 |
| resp | nresp_dbs_bilat | | 30 | 0.184 | 0.220 | 0.010 | 0.014 | 1.29E-02 | 7.28E-31 |
| hc | resp_dbs_off | | 15 | 0.203 | 0.184 | 0.001 | 0.004 | 5.93E-07 | 0.00E+00 |
| hc | resp_dbs_off | | 30 | 0.202 | 0.182 | 0.002 | 0.005 | 1.63E-04 | 9.95E-268 |
| hc | nresp_dbs_off | | 15 | 0.203 | 0.180 | 0.001 | 0.004 | 2.72E-09 | 0.00E+00 |
| hc | nresp_dbs_off | | 30 | 0.202 | 0.180 | 0.002 | 0.005 | 2.88E-05 | 5.84E-264 |
| resp | nresp_dbs_off | | 15 | 0.180 | 0.184 | 0.004 | 0.006 | 4.56E-01 | 1.25E-103 |
| resp | nresp_dbs_off | | 30 | 0.180 | 0.182 | 0.005 | 0.007 | 7.79E-01 | 2.39E-57 |
| caudalmiddlefrontal – left hemisphere – gamma band | | | | | | | | | |
| Compared factor phenotypes | | | epoch size [s] | values | | variance | | significance | |
| phenotype A | phenotype B | | - | phen. A | phen. B | phen. A | phen. B | phen. A | intercept |
| resp_dbs_off | resp_dbs_bilat | | 15 | 0.187 | 0.223 | 0.018 | 0.007 | 6.64E-08 | 2.21E-26 |
| resp_dbs_off | resp_dbs_bilat | | 30 | 0.187 | 0.222 | 0.018 | 0.009 | 7.72E-05 | 2.84E-25 |
| nresp_dbs_off | nresp_dbs_bilat | | 15 | 0.171 | 0.174 | 0.015 | 0.003 | 4.38E-01 | 1.26E-28 |
| nresp_dbs_off | nresp_dbs_bilat | | 30 | 0.171 | 0.172 | 0.015 | 0.004 | 7.18E-01 | 1.54E-28 |
| hc | resp_dbs_bilat | | 15 | 0.198 | 0.223 | 0.002 | 0.005 | 1.34E-06 | 0.00E+00 |
| hc | resp_dbs_bilat | | 30 | 0.197 | 0.222 | 0.002 | 0.007 | 5.74E-04 | 2.88E-222 |
| hc | nresp_dbs_bilat | | 15 | 0.198 | 0.175 | 0.002 | 0.004 | 8.62E-08 | 0.00E+00 |
| hc | nresp_dbs_bilat | | 30 | 0.197 | 0.173 | 0.002 | 0.006 | 4.35E-05 | 2.95E-246 |
| resp | nresp_dbs_bilat | | 15 | 0.175 | 0.223 | 0.007 | 0.010 | 3.32E-06 | 1.26E-56 |
| resp | nresp_dbs_bilat | | 30 | 0.173 | 0.222 | 0.010 | 0.014 | 7.27E-04 | 1.27E-29 |
| hc | resp_dbs_off | | 15 | 0.198 | 0.184 | 0.001 | 0.004 | 4.67E-04 | 0.00E+00 |
| hc | resp_dbs_off | | 30 | 0.197 | 0.184 | 0.002 | 0.005 | 1.14E-02 | 1.92E-255 |
| hc | nresp_dbs_off | | 15 | 0.198 | 0.172 | 0.002 | 0.004 | 1.21E-10 | 0.00E+00 |
| hc | nresp_dbs_off | | 30 | 0.197 | 0.171 | 0.002 | 0.005 | 2.11E-06 | 1.89E-248 |
| resp | nresp_dbs_off | | 15 | 0.172 | 0.184 | 0.004 | 0.006 | 3.60E-02 | 2.83E-97 |
| resp | nresp_dbs_off | | 30 | 0.171 | 0.184 | 0.005 | 0.008 | 1.15E-01 | 5.61E-52 |
| retrosplenial – left hemisphere – gamma band | | | | | | | | | |
| Compared factor phenotypes | | | epoch size [s] | values | | variance | | significance | |
| phenotype A | phenotype B | | - | phen. A | phen. B | phen. A | phen. B | phen. A | intercept |
| resp_dbs_off | resp_dbs_bilat | | 15 | 0.184 | 0.219 | 0.026 | 0.007 | 1.53E-06 | 7.80E-13 |
| resp_dbs_off | resp_dbs_bilat | | 30 | 0.183 | 0.221 | 0.027 | 0.011 | 4.44E-04 | 6.50E-12 |
| nresp_dbs_off | nresp_dbs_bilat | | 15 | 0.191 | 0.196 | 0.014 | 0.003 | 9.14E-02 | 9.93E-41 |
| nresp_dbs_off | nresp_dbs_bilat | | 30 | 0.190 | 0.196 | 0.014 | 0.003 | 9.30E-02 | 1.49E-41 |
| hc | resp_dbs_bilat | | 15 | 0.206 | 0.218 | 0.002 | 0.006 | 3.61E-02 | 0.00E+00 |
| hc | resp_dbs_bilat | | 30 | 0.204 | 0.221 | 0.003 | 0.008 | 4.30E-02 | 3.46E-206 |
| hc | nresp_dbs_bilat | | 15 | 0.206 | 0.193 | 0.001 | 0.004 | 2.38E-03 | 0.00E+00 |
| hc | nresp_dbs_bilat | | 30 | 0.204 | 0.192 | 0.002 | 0.006 | 3.29E-02 | 1.80E-258 |
| resp | nresp_dbs_bilat | | 15 | 0.193 | 0.218 | 0.009 | 0.013 | 4.98E-02 | 6.03E-49 |
| resp | nresp_dbs_bilat | | 30 | 0.192 | 0.221 | 0.013 | 0.018 | 1.18E-01 | 2.46E-25 |

| | | | | | | | | | |
|---|---|---|---|---|---|---|---|---|---|
| hc | resp_dbs_off | | 15 | 0.206 | 0.181 | 0.001 | 0.004 | 1.58E-09 | 0.00E+00 |
| hc | resp_dbs_off | | 30 | 0.204 | 0.180 | 0.002 | 0.005 | 5.04E-06 | 1.32E-263 |
| hc | nresp_dbs_off | | 15 | 0.206 | 0.192 | 0.001 | 0.004 | 7.57E-04 | 0.00E+00 |
| hc | nresp_dbs_off | | 30 | 0.204 | 0.190 | 0.002 | 0.005 | 6.60E-03 | 1.91E-262 |
| resp | nresp_dbs_off | | 15 | 0.192 | 0.181 | 0.004 | 0.006 | 6.35E-02 | 1.72E-103 |
| resp | nresp_dbs_off | | 30 | 0.190 | 0.180 | 0.005 | 0.008 | 1.80E-01 | 9.86E-58 |
| lingual – left hemisphere – gamma band | | | | | | | | | |
| Compared factor phenotypes | | | epoch size [s] | values | | variance | | significance | |
| phenotype A | phenotype B | | - | phen. A | phen. B | phen. A | phen. B | phen. A | intercept |
| resp_dbs_off | resp_dbs_bilat | | 15 | 0.186 | 0.200 | 0.019 | 0.005 | 2.42E-03 | 1.47E-23 |
| resp_dbs_off | resp_dbs_bilat | | 30 | 0.184 | 0.199 | 0.020 | 0.006 | 9.83E-03 | 6.54E-21 |
| nresp_dbs_off | nresp_dbs_bilat | | 15 | 0.180 | 0.190 | 0.014 | 0.002 | 5.16E-05 | 1.53E-39 |
| nresp_dbs_off | nresp_dbs_bilat | | 30 | 0.179 | 0.188 | 0.014 | 0.003 | 6.91E-03 | 3.65E-40 |
| hc | resp_dbs_bilat | | 15 | 0.200 | 0.199 | 0.002 | 0.005 | 6.92E-01 | 0.00E+00 |
| hc | resp_dbs_bilat | | 30 | 0.199 | 0.199 | 0.002 | 0.006 | 9.83E-01 | 3.87E-238 |
| hc | nresp_dbs_bilat | | 15 | 0.200 | 0.187 | 0.001 | 0.004 | 1.62E-03 | 0.00E+00 |
| hc | nresp_dbs_bilat | | 30 | 0.199 | 0.187 | 0.002 | 0.005 | 2.71E-02 | 9.36E-258 |
| resp | nresp_dbs_bilat | | 15 | 0.187 | 0.199 | 0.006 | 0.008 | 1.80E-01 | 9.30E-70 |
| resp | nresp_dbs_bilat | | 30 | 0.187 | 0.199 | 0.008 | 0.012 | 3.07E-01 | 2.85E-36 |
| hc | resp_dbs_off | | 15 | 0.200 | 0.182 | 0.001 | 0.004 | 3.36E-06 | 0.00E+00 |
| hc | resp_dbs_off | | 30 | 0.199 | 0.180 | 0.002 | 0.005 | 3.22E-04 | 3.35E-258 |
| hc | nresp_dbs_off | | 15 | 0.200 | 0.180 | 0.001 | 0.004 | 5.10E-08 | 0.00E+00 |
| hc | nresp_dbs_off | | 30 | 0.199 | 0.179 | 0.002 | 0.005 | 5.94E-05 | 7.36E-263 |
| resp | nresp_dbs_off | | 15 | 0.180 | 0.182 | 0.004 | 0.006 | 7.39E-01 | 3.51E-101 |
| resp | nresp_dbs_off | | 30 | 0.179 | 0.180 | 0.006 | 0.008 | 9.03E-01 | 2.94E-53 |
| paracentral – left hemisphere – gamma band | | | | | | | | | |
| Compared factor phenotypes | | | epoch size [s] | values | | variance | | significance | |
| phenotype A | phenotype B | | - | phen. A | phen. B | phen. A | phen. B | phen. A | intercept |
| resp_dbs_off | resp_dbs_bilat | | 15 | 0.179 | 0.200 | 0.020 | 0.005 | 9.85E-06 | 2.89E-19 |
| resp_dbs_off | resp_dbs_bilat | | 30 | 0.177 | 0.198 | 0.020 | 0.006 | 3.50E-04 | 8.47E-19 |
| nresp_dbs_off | nresp_dbs_bilat | | 15 | 0.185 | 0.189 | 0.012 | 0.003 | 1.87E-01 | 2.80E-53 |
| nresp_dbs_off | nresp_dbs_bilat | | 30 | 0.183 | 0.188 | 0.012 | 0.004 | 2.20E-01 | 2.41E-55 |
| hc | resp_dbs_bilat | | 15 | 0.204 | 0.200 | 0.002 | 0.005 | 4.33E-01 | 0.00E+00 |
| hc | resp_dbs_bilat | | 30 | 0.202 | 0.198 | 0.002 | 0.006 | 6.20E-01 | 1.14E-239 |
| hc | nresp_dbs_bilat | | 15 | 0.204 | 0.190 | 0.001 | 0.004 | 5.26E-04 | 0.00E+00 |
| hc | nresp_dbs_bilat | | 30 | 0.202 | 0.189 | 0.002 | 0.005 | 1.61E-02 | 1.52E-263 |
| resp | nresp_dbs_bilat | | 15 | 0.190 | 0.200 | 0.006 | 0.009 | 2.47E-01 | 8.17E-70 |
| resp | nresp_dbs_bilat | | 30 | 0.189 | 0.198 | 0.008 | 0.012 | 4.10E-01 | 4.71E-37 |
| hc | resp_dbs_off | | 15 | 0.204 | 0.174 | 0.001 | 0.004 | 1.36E-13 | 0.00E+00 |
| hc | resp_dbs_off | | 30 | 0.202 | 0.174 | 0.002 | 0.005 | 2.23E-07 | 2.21E-261 |
| hc | nresp_dbs_off | | 15 | 0.204 | 0.186 | 0.001 | 0.004 | 9.27E-07 | 0.00E+00 |
| hc | nresp_dbs_off | | 30 | 0.202 | 0.184 | 0.002 | 0.005 | 4.97E-04 | 3.59E-267 |
| resp | nresp_dbs_off | | 15 | 0.186 | 0.174 | 0.004 | 0.006 | 5.49E-02 | 3.51E-104 |
| resp | nresp_dbs_off | | 30 | 0.184 | 0.174 | 0.005 | 0.008 | 1.80E-01 | 1.29E-55 |
| opercularis – left hemisphere – gamma band | | | | | | | | | |
| Compared factor phenotypes | | | epoch size [s] | values | | variance | | significance | |
| phenotype A | phenotype B | | - | phen. A | phen. B | phen. A | phen. B | phen. A | intercept |
| resp_dbs_off | resp_dbs_bilat | | 15 | 0.185 | 0.196 | 0.010 | 0.003 | 1.95E-03 | 3.35E-74 |
| resp_dbs_off | resp_dbs_bilat | | 30 | 0.183 | 0.194 | 0.011 | 0.004 | 1.45E-02 | 6.52E-67 |
| nresp_dbs_off | nresp_dbs_bilat | | 15 | 0.182 | 0.181 | 0.017 | 0.003 | 8.78E-01 | 2.60E-25 |
| nresp_dbs_off | nresp_dbs_bilat | | 30 | 0.181 | 0.182 | 0.017 | 0.004 | 8.82E-01 | 2.53E-25 |
| hc | resp_dbs_bilat | | 15 | 0.199 | 0.196 | 0.001 | 0.004 | 5.03E-01 | 0.00E+00 |
| hc | resp_dbs_bilat | | 30 | 0.197 | 0.194 | 0.002 | 0.005 | 4.85E-01 | 4.11E-265 |
| hc | nresp_dbs_bilat | | 15 | 0.199 | 0.181 | 0.001 | 0.004 | 1.88E-05 | 0.00E+00 |
| hc | nresp_dbs_bilat | | 30 | 0.197 | 0.180 | 0.002 | 0.006 | 2.55E-03 | 2.49E-252 |
| resp | nresp_dbs_bilat | | 15 | 0.181 | 0.196 | 0.005 | 0.007 | 2.29E-02 | 3.17E-81 |

| | | | | | | | | | |
|---|---|---|---|---|---|---|---|---|---|
| resp | nresp_dbs_bilat | | 30 | 0.180 | 0.194 | 0.007 | 0.009 | 1.50E-01 | 6.57E-43 |
| hc | resp_dbs_off | | 15 | 0.199 | 0.184 | 0.001 | 0.004 | 5.87E-05 | 0.00E+00 |
| hc | resp_dbs_off | | 30 | 0.197 | 0.181 | 0.002 | 0.005 | 1.34E-03 | 2.28E-267 |
| hc | nresp_dbs_off | | 15 | 0.199 | 0.182 | 0.001 | 0.004 | 1.09E-05 | 0.00E+00 |
| hc | nresp_dbs_off | | 30 | 0.197 | 0.182 | 0.002 | 0.005 | 3.51E-03 | 1.30E-257 |
| resp | nresp_dbs_off | | 15 | 0.182 | 0.184 | 0.004 | 0.006 | 7.51E-01 | 1.73E-98 |
| resp | nresp_dbs_off | | 30 | 0.182 | 0.181 | 0.006 | 0.008 | 9.14E-01 | 1.50E-52 |
| triangularis – left hemisphere – gamma band | | | | | | | | | |
| Compared factor phenotypes | | | epoch size [s] | values | | variance | | significance | |
| phenotype A | phenotype B | | - | phen. A | phen. B | phen. A | phen. B | phen. A | intercept |
| resp_dbs_off | resp_dbs_bilat | | 15 | 0.189 | 0.201 | 0.012 | 0.003 | 3.03E-05 | 7.73E-55 |
| resp_dbs_off | resp_dbs_bilat | | 30 | 0.188 | 0.201 | 0.012 | 0.004 | 6.82E-04 | 2.62E-55 |
| nresp_dbs_off | nresp_dbs_bilat | | 15 | 0.185 | 0.190 | 0.016 | 0.002 | 5.50E-02 | 1.31E-30 |
| nresp_dbs_off | nresp_dbs_bilat | | 30 | 0.185 | 0.188 | 0.016 | 0.003 | 2.50E-01 | 4.07E-31 |
| hc | resp_dbs_bilat | | 15 | 0.205 | 0.201 | 0.001 | 0.004 | 3.27E-01 | 0.00E+00 |
| hc | resp_dbs_bilat | | 30 | 0.204 | 0.201 | 0.002 | 0.005 | 6.11E-01 | 3.82E-271 |
| hc | nresp_dbs_bilat | | 15 | 0.205 | 0.189 | 0.001 | 0.004 | 2.50E-04 | 0.00E+00 |
| hc | nresp_dbs_bilat | | 30 | 0.204 | 0.188 | 0.002 | 0.005 | 3.77E-03 | 2.07E-261 |
| resp | nresp_dbs_bilat | | 15 | 0.189 | 0.201 | 0.005 | 0.006 | 7.85E-02 | 7.08E-87 |
| resp | nresp_dbs_bilat | | 30 | 0.188 | 0.201 | 0.006 | 0.009 | 1.33E-01 | 3.44E-46 |
| hc | resp_dbs_off | | 15 | 0.205 | 0.186 | 0.001 | 0.004 | 6.98E-07 | 0.00E+00 |
| hc | resp_dbs_off | | 30 | 0.204 | 0.185 | 0.002 | 0.005 | 1.81E-04 | 8.30E-271 |
| hc | nresp_dbs_off | | 15 | 0.205 | 0.186 | 0.001 | 0.004 | 6.20E-07 | 0.00E+00 |
| hc | nresp_dbs_off | | 30 | 0.204 | 0.186 | 0.002 | 0.005 | 4.64E-04 | 2.35E-267 |
| resp | nresp_dbs_off | | 15 | 0.186 | 0.186 | 0.004 | 0.006 | 9.81E-01 | 1.21E-100 |
| resp | nresp_dbs_off | | 30 | 0.186 | 0.185 | 0.006 | 0.008 | 8.69E-01 | 2.94E-54 |
| postcentral – left hemisphere – gamma band | | | | | | | | | |
| Compared factor phenotypes | | | epoch size [s] | values | | variance | | significance | |
| phenotype A | phenotype B | | - | phen. A | phen. B | phen. A | phen. B | phen. A | intercept |
| resp_dbs_off | resp_dbs_bilat | | 15 | 0.181 | 0.228 | 0.022 | 0.007 | 9.00E-11 | 4.71E-16 |
| resp_dbs_off | resp_dbs_bilat | | 30 | 0.179 | 0.226 | 0.023 | 0.010 | 2.33E-06 | 1.92E-15 |
| nresp_dbs_off | nresp_dbs_bilat | | 15 | 0.176 | 0.180 | 0.017 | 0.003 | 1.23E-01 | 8.45E-26 |
| nresp_dbs_off | nresp_dbs_bilat | | 30 | 0.174 | 0.179 | 0.017 | 0.004 | 2.27E-01 | 2.20E-25 |
| hc | resp_dbs_bilat | | 15 | 0.197 | 0.229 | 0.002 | 0.006 | 2.75E-08 | 0.00E+00 |
| hc | resp_dbs_bilat | | 30 | 0.195 | 0.226 | 0.003 | 0.008 | 6.19E-05 | 3.07E-205 |
| hc | nresp_dbs_bilat | | 15 | 0.197 | 0.181 | 0.002 | 0.005 | 5.73E-04 | 0.00E+00 |
| hc | nresp_dbs_bilat | | 30 | 0.195 | 0.180 | 0.002 | 0.006 | 2.21E-02 | 4.65E-236 |
| resp | nresp_dbs_bilat | | 15 | 0.181 | 0.229 | 0.009 | 0.012 | 1.19E-04 | 1.07E-47 |
| resp | nresp_dbs_bilat | | 30 | 0.180 | 0.226 | 0.012 | 0.017 | 7.57E-03 | 1.54E-25 |
| hc | resp_dbs_off | | 15 | 0.197 | 0.179 | 0.002 | 0.004 | 3.28E-05 | 0.00E+00 |
| hc | resp_dbs_off | | 30 | 0.195 | 0.176 | 0.002 | 0.006 | 9.35E-04 | 1.28E-248 |
| hc | nresp_dbs_off | | 15 | 0.197 | 0.176 | 0.002 | 0.004 | 1.95E-06 | 0.00E+00 |
| hc | nresp_dbs_off | | 30 | 0.195 | 0.176 | 0.002 | 0.006 | 9.04E-04 | 1.81E-243 |
| resp | nresp_dbs_off | | 15 | 0.176 | 0.179 | 0.004 | 0.006 | 6.74E-01 | 5.94E-98 |
| resp | nresp_dbs_off | | 30 | 0.176 | 0.176 | 0.005 | 0.008 | 9.88E-01 | 3.82E-53 |
| posteriorcingulat – left hemisphere – gamma band | | | | | | | | | |
| compared factors | | | epoch size [s] | values | | variance | | significance | |
| phenotype A | phenotype B | | - | phen. A | phen. B | phen. A | phen. B | phen. A | intercept |
| resp_dbs_off | resp_dbs_bilat | | 15 | 0.183 | 0.202 | 0.019 | 0.005 | 1.13E-04 | 3.23E-21 |
| resp_dbs_off | resp_dbs_bilat | | 30 | 0.180 | 0.200 | 0.019 | 0.006 | 1.30E-03 | 1.08E-20 |
| nresp_dbs_off | nresp_dbs_bilat | | 15 | 0.191 | 0.190 | 0.010 | 0.003 | 6.70E-01 | 3.15E-74 |
| nresp_dbs_off | nresp_dbs_bilat | | 30 | 0.191 | 0.188 | 0.010 | 0.004 | 5.76E-01 | 6.83E-77 |
| hc | resp_dbs_bilat | | 15 | 0.204 | 0.202 | 0.002 | 0.004 | 6.52E-01 | 0.00E+00 |
| hc | resp_dbs_bilat | | 30 | 0.202 | 0.200 | 0.002 | 0.006 | 7.75E-01 | 1.09E-244 |
| hc | nresp_dbs_bilat | | 15 | 0.204 | 0.191 | 0.001 | 0.004 | 8.10E-04 | 0.00E+00 |
| hc | nresp_dbs_bilat | | 30 | 0.202 | 0.190 | 0.002 | 0.005 | 1.64E-02 | 3.68E-270 |

| | | | | | | | | | |
|---|---|---|---|---|---|---|---|---|---|
| resp | nresp_dbs_bilat | | 15 | 0.191 | 0.202 | 0.006 | 0.008 | 2.01E-01 | 2.35E-72 |
| resp | nresp_dbs_bilat | | 30 | 0.190 | 0.200 | 0.008 | 0.012 | 3.64E-01 | 8.90E-38 |
| hc | resp_dbs_off | | 15 | 0.204 | 0.178 | 0.001 | 0.004 | 1.38E-11 | 0.00E+00 |
| hc | resp_dbs_off | | 30 | 0.202 | 0.176 | 0.002 | 0.005 | 7.73E-07 | 2.52E-267 |
| hc | nresp_dbs_off | | 15 | 0.204 | 0.191 | 0.001 | 0.004 | 2.73E-04 | 0.00E+00 |
| hc | nresp_dbs_off | | 30 | 0.202 | 0.191 | 0.002 | 0.005 | 1.98E-02 | 7.04E-276 |
| resp | nresp_dbs_off | | 15 | 0.191 | 0.178 | 0.004 | 0.006 | 1.87E-02 | 5.02E-108 |
| resp | nresp_dbs_off | | 30 | 0.191 | 0.176 | 0.005 | 0.007 | 5.06E-02 | 3.04E-59 |
| precentral – left hemisphere – gamma band | | | | | | | | | |
| compared factors | | | epoch size [s] | values | | variance | | significance | |
| phenotype A | phenotype B | | - | phen. A | phen. B | phen. A | phen. B | phen. A | intercept |
| resp_dbs_off | resp_dbs_bilat | | 15 | 0.177 | 0.224 | 0.021 | 0.007 | 1.48E-12 | 1.89E-17 |
| resp_dbs_off | resp_dbs_bilat | | 30 | 0.176 | 0.222 | 0.021 | 0.009 | 5.88E-07 | 4.06E-17 |
| nresp_dbs_off | nresp_dbs_bilat | | 15 | 0.174 | 0.179 | 0.017 | 0.003 | 6.61E-02 | 1.21E-25 |
| nresp_dbs_off | nresp_dbs_bilat | | 30 | 0.173 | 0.177 | 0.016 | 0.003 | 2.17E-01 | 4.39E-26 |
| hc | resp_dbs_bilat | | 15 | 0.197 | 0.224 | 0.002 | 0.006 | 1.70E-06 | 0.00E+00 |
| hc | resp_dbs_bilat | | 30 | 0.195 | 0.222 | 0.003 | 0.008 | 5.99E-04 | 7.55E-206 |
| hc | nresp_dbs_bilat | | 15 | 0.197 | 0.180 | 0.002 | 0.005 | 6.58E-04 | 0.00E+00 |
| hc | nresp_dbs_bilat | | 30 | 0.195 | 0.178 | 0.002 | 0.007 | 1.32E-02 | 3.57E-229 |
| resp | nresp_dbs_bilat | | 15 | 0.180 | 0.224 | 0.008 | 0.011 | 1.40E-04 | 5.99E-51 |
| resp | nresp_dbs_bilat | | 30 | 0.178 | 0.222 | 0.011 | 0.015 | 6.49E-03 | 1.98E-27 |
| hc | resp_dbs_off | | 15 | 0.197 | 0.175 | 0.002 | 0.004 | 7.22E-07 | 0.00E+00 |
| hc | resp_dbs_off | | 30 | 0.195 | 0.174 | 0.002 | 0.006 | 5.36E-04 | 1.79E-239 |
| hc | nresp_dbs_off | | 15 | 0.197 | 0.176 | 0.002 | 0.004 | 2.40E-06 | 0.00E+00 |
| hc | nresp_dbs_off | | 30 | 0.195 | 0.175 | 0.002 | 0.006 | 7.71E-04 | 1.39E-235 |
| resp | nresp_dbs_off | | 15 | 0.176 | 0.175 | 0.004 | 0.006 | 8.57E-01 | 1.61E-98 |
| resp | nresp_dbs_off | | 30 | 0.175 | 0.174 | 0.005 | 0.008 | 9.28E-01 | 3.05E-53 |
| rostralmiddlefrontal – left hemisphere – gamma band | | | | | | | | | |
| compared factors | | | epoch size [s] | values | | variance | | significance | |
| phenotype A | phenotype B | | - | phen. A | phen. B | phen. A | phen. B | phen. A | intercept |
| resp_dbs_off | resp_dbs_bilat | | 15 | 0.187 | 0.204 | 0.014 | 0.004 | 1.02E-05 | 3.35E-41 |
| resp_dbs_off | resp_dbs_bilat | | 30 | 0.186 | 0.203 | 0.014 | 0.005 | 4.19E-04 | 9.93E-41 |
| nresp_dbs_off | nresp_dbs_bilat | | 15 | 0.173 | 0.183 | 0.013 | 0.003 | 6.60E-04 | 4.69E-43 |
| nresp_dbs_off | nresp_dbs_bilat | | 30 | 0.173 | 0.180 | 0.012 | 0.004 | 4.93E-02 | 8.68E-44 |
| hc | resp_dbs_bilat | | 15 | 0.200 | 0.204 | 0.001 | 0.004 | 2.57E-01 | 0.00E+00 |
| hc | resp_dbs_bilat | | 30 | 0.199 | 0.203 | 0.002 | 0.005 | 3.93E-01 | 4.40E-261 |
| hc | nresp_dbs_bilat | | 15 | 0.200 | 0.183 | 0.001 | 0.004 | 4.37E-05 | 0.00E+00 |
| hc | nresp_dbs_bilat | | 30 | 0.199 | 0.182 | 0.002 | 0.005 | 1.59E-03 | 6.10E-265 |
| resp | nresp_dbs_bilat | | 15 | 0.183 | 0.204 | 0.005 | 0.007 | 1.79E-03 | 2.55E-84 |
| resp | nresp_dbs_bilat | | 30 | 0.182 | 0.203 | 0.006 | 0.009 | 1.81E-02 | 2.56E-45 |
| hc | resp_dbs_off | | 15 | 0.200 | 0.183 | 0.001 | 0.004 | 1.35E-05 | 0.00E+00 |
| hc | resp_dbs_off | | 30 | 0.199 | 0.182 | 0.002 | 0.005 | 1.09E-03 | 1.76E-265 |
| hc | nresp_dbs_off | | 15 | 0.200 | 0.173 | 0.001 | 0.004 | 3.42E-13 | 0.00E+00 |
| hc | nresp_dbs_off | | 30 | 0.199 | 0.173 | 0.002 | 0.005 | 9.15E-08 | 1.40E-269 |
| resp | nresp_dbs_off | | 15 | 0.173 | 0.183 | 0.004 | 0.006 | 8.06E-02 | 4.44E-97 |
| resp | nresp_dbs_off | | 30 | 0.173 | 0.182 | 0.005 | 0.008 | 2.28E-01 | 1.15E-53 |
| supramarginal – left hemisphere – gamma band | | | | | | | | | |
| compared factors | | | epoch size [s] | values | | variance | | significance | |
| phenotype A | phenotype B | | - | phen. A | phen. B | phen. A | phen. B | phen. A | intercept |
| resp_dbs_off | resp_dbs_bilat | | 15 | 0.184 | 0.231 | 0.024 | 0.008 | 2.30E-09 | 4.28E-14 |
| resp_dbs_off | resp_dbs_bilat | | 30 | 0.181 | 0.228 | 0.025 | 0.011 | 1.83E-05 | 2.37E-13 |
| nresp_dbs_off | nresp_dbs_bilat | | 15 | 0.176 | 0.181 | 0.015 | 0.003 | 5.42E-02 | 3.52E-30 |
| nresp_dbs_off | nresp_dbs_bilat | | 30 | 0.176 | 0.180 | 0.015 | 0.004 | 2.37E-01 | 2.58E-30 |
| hc | resp_dbs_bilat | | 15 | 0.199 | 0.231 | 0.002 | 0.006 | 5.71E-08 | 0.00E+00 |
| hc | resp_dbs_bilat | | 30 | 0.197 | 0.228 | 0.003 | 0.008 | 1.46E-04 | 5.99E-204 |
| hc | nresp_dbs_bilat | | 15 | 0.199 | 0.182 | 0.002 | 0.004 | 4.70E-05 | 0.00E+00 |

| | | | | | | | | | |
|---|---|---|---|---|---|---|---|---|---|
| hc | nresp_dbs_bilat | | 30 | 0.197 | 0.181 | 0.002 | 0.006 | 7.36E-03 | 5.48E-247 |
| resp | nresp_dbs_bilat | | 15 | 0.182 | 0.231 | 0.009 | 0.013 | 2.01E-04 | 7.42E-45 |
| resp | nresp_dbs_bilat | | 30 | 0.181 | 0.228 | 0.013 | 0.018 | 1.13E-02 | 4.87E-24 |
| hc | resp_dbs_off | | 15 | 0.199 | 0.180 | 0.001 | 0.004 | 6.40E-07 | 0.00E+00 |
| hc | resp_dbs_off | | 30 | 0.197 | 0.177 | 0.002 | 0.005 | 2.20E-04 | 1.34E-257 |
| hc | nresp_dbs_off | | 15 | 0.199 | 0.178 | 0.001 | 0.004 | 1.94E-08 | 0.00E+00 |
| hc | nresp_dbs_off | | 30 | 0.197 | 0.177 | 0.002 | 0.005 | 1.04E-04 | 1.70E-255 |
| resp | nresp_dbs_off | | 15 | 0.178 | 0.180 | 0.004 | 0.006 | 7.06E-01 | 7.28E-103 |
| resp | nresp_dbs_off | | 30 | 0.177 | 0.177 | 0.005 | 0.008 | 9.29E-01 | 5.45E-55 |
